# Implementation of Human-in-the-Loop ChatGPT-based Patient Screening Across Multiple Diverse Clinical Trials

**DOI:** 10.64898/2026.03.20.26348890

**Authors:** Michael Dohopolski, Kate Esselink, Neil Desai, Bailey Grones, Toral Patel, Steve Jiang, Eric Peterson, Ann Marie Navar

## Abstract

**Purpose:** Manual screening for trial eligibility is inefficient and costly. We prospectively evaluated a large language model (LLM)-assisted prescreening workflow across multiple active trials.

**Methods:** We deployed a retrieval-augmented generation LLM-based pipeline across multiple trials at an academic medical center. Structured electronic health record data and free-text notes were used by the LLM to classify each criterion as either *met, likely met, likely not met, not met, uncertain*, or *no documentation found* with accompanying rationale. Coordinators were given a sorted patient list based on the LLM-based eligibility to be reviewed and provide their assessment of each criterion, and final prescreening status (success vs failure). Criterion-level performance– accuracy, sensitivity, specificity, positive predictive value (PPV), negative predictive value (NPV), and F1 score–was calculated and measured over time. Patient prescreening status was evaluated as a function of percent of individual AI criteria met (60-80% and ≥ **80**%).

**Results:** From 10/2024–9/2025, 39,182 patients were prescreened by our LLM-workflow across 26 (21 oncology and 5 non-oncology) studies with 112 distinct criteria; 914 patients with high likelihood for eligibility underwent coordinator review (total 5,096 criteria). Aggregated individual criterion-level performance: accuracy 0.94 (95% CI, 0.92–0.96), sensitivity 0.98 (0.97–0.99), specificity 0.81 (0.71–0.88), PPV 0.95 (0.92–0.97), NPV 0.93 (0.90–0.95), F1 0.97 (0.95–0.97). Twenty-seven criteria prompts across 14/26 trials were automatically updated based on coordinator feedback. Patients were more likely to be reviewed by coordinators (544/987, 55.1% vs 372/397, 93.7%) and more likely to be labeled as a prescreen success (104/544, 19.1% vs 162/372, 43.5%) when ≥ **80%** AI-labeled criteria were *met* or *likely met* vs 60-80%. Cost averaged $0.12 per patient.

**Conclusion:** A LLM-assisted, human-in-the-loop prescreening workflow achieved high criterion-level performance at low cost across a variety of actively enrolling clinical trials. Structured coordinator feedback supported an automated learning workflow, enhancing screening operations while maintaining human oversight.

## Background

Clinical trials are essential for validating new medical treatments, but often face challenges with patient enrollment. Insufficient accrual can significantly delay research, limit the predictive power of results, and inflate costs [1–3]. Manual patient eligibility screening exacerbates these challenges by being time-consuming and resource-intensive, highlighting the need for efficient automation [4, 5].

Recently, large language models (LLMs) have shown promise for automating clinical trial eligibility screening using advanced natural language processing. Unlike earlier natural language processing approaches that required extensive task-specific customization [6–9], LLMs offer broader applicability with fewer engineering constraints. Early studies demonstrate feasibility in real-world or realistic settings across diverse designs and disease areas [10–13]. To date, however, reports of using LLMs for prospective trial screening have been limited to individual trials in specific clinical areas, while their generalizability across diverse trial designs remains largely unevaluated. Additionally, LLM pre-screening has not been robustly integrated with real-time human-in-the-loop learning strategies that may facilitate earlier refinement and acceptance.

In this study, we describe how LLM-enabled prescreening was operationalized across multiple concurrent trials at a large academic medical center. We also assessed the overall performance and cost of LLMs used to identify patients potentially eligible for clinical trials. Our implementation emphasizes structured human oversight and a strategy to iteratively improve the LLM algorithms by incorporating human reviewer feedback, with the goal of providing a pragmatic template for integrating LLMs into routine prescreening workflows.

## Methods

### Onboarding clinical trials into artificial intelligence (AI) workflow

The LLM/AI-enabled prescreening workflow was made available to study teams within our institution starting in October 2024. For each participating study, the principal investigator (PI) amended the trial-specific Institutional Review Board (IRB)-approved protocol to add AI-team members as study personnel. The AI team configured and maintained the prescreening pipeline and reviewed performance patterns with study staff/coordinators, while all approach, consent, and enrollment decisions remained the responsibility of the trial team. At onboarding, each study team: (i) selected 2– 7 trial-specific eligibility criteria and ranked the criteria to be used in a tiered workflow: a single main criterion, which we named Tier 1 criterion, determined whether remaining criteria (all designated as Tier 2) were evaluated; (ii) identified the providers or clinics to screen; (iii) confirmed structured and unstructured text to be reviewed (encounters/clinical notes, pathology reports, imaging reports, lab reports); and (iv) agreed to brief feedback sessions to address discordance and edge cases. A standardized setup form captured study-level parameters for each trial (Supplement B: Standardized Setup Form).

Each eligibility criterion text was automatically transformed into a tailored prompt with three elements: the criterion name, verbatim protocol text, and a concise *tips* section that mapped logic to standardized labels (*met, likely met, likely not met, not met, uncertain, no documentation found* ). Additional medical context (synonyms, definitions, etc) was added from PubMed summaries generated from our PubMed summary agent to improve accuracy[13, 14]. Further details can be found in Supplement B: Generating eligibility prompts.

### Patient data and preprocessing

The electronic health record (EHR) came from Epic (Epic Systems Corporation, Verona, WI). Data was extracted from Caboodle and Clarity, which are updated nightly. To accommodate this latency, prescreen patient lists were generated prospectively several times a week with a short look-ahead window (typically 2-4 days). These lists were created from upcoming schedules for providers or clinics that the study teams wanted to be screened. For each patient, clinical text within a trial-specific window (typically 0–180 days) was retrieved across encounters, imaging reports (CT, MRI, PET, etc), pathology reports, and laboratory results; scanned documents (e.g. external PDFs) were excluded per institutional policy. Where appropriate, filters were applied by specialty, note type, specific laboratory tests, and date of documentation; imaging and pathology were typically filtered only by date of documentation. Medical texts were transformed into a per-patient vector database to enable similarity search via a retrieval-augmented generation-based approach. A hierarchical node parser was used to create multi-resolution nodes (practically mimicking document → paragraph → sentence).

### AI prescreening

Criterion evaluation followed a tiered workflow. First, the Tier 1 criterion was assessed; the pipeline proceeded to Tier 2 criteria only if Tier 1 was labeled by AI as *met* or *likely met*. If Tier 1 was labeled as *likely not met, not met, uncertain*, or *no documentation found*, then Tier 2 criteria were not evaluated. For each criterion evaluated, the pipeline selected the top 30 most relevant nodes and provided them to the LLM. The LLM returned (a) a status label (*met, likely met, likely not met, not met, uncertain, no documentation found* ); (b) a descriptive 5-point certainty; and (c) a brief rationale referencing the specific clinical evidence (examples in **Table S1**; Supplement A: Example criteria and AI outputs). All evaluations were performed in a HIPAA-compliant Microsoft Azure environment. The base LLM changed as newer and/or lower cost models became available: GPT-4o (October 2024–March 2025), GPT-4o-mini (March 2025–June 2025), and GPT-4.1-mini (June 2025–present). Embeddings for retrieval were generated with `text-embedding-3-large`.

### Trial coordinator review and AI refinement

Patients who exceeded a threshold for the proportion of criteria AI labeled *met* or *likely met* were displayed in a sorted, coordinator-facing dashboard that showed results from both Tier 1 and Tier 2 criteria for review. Further details were provided when coordinators accessed a link to a particular patient, which showed the AI-labeled status and rationale, the level of confidence reported by the LLM, and the specific clinical data supporting the result, including the location of those data within the medical record (e.g., note ID, document date). Coordinators were asked to at least review patients who met≥50% of the criteria, while all other patients were left for optional review. After the first 1-3 months, as criterion-specific performance improved, coordinators could restrict review to patients who AI deemed to meet a greater proportion of criteria (ie, ≥ 75%). Again, no restrictions were in place to limit the review of any other patients. As mentioned previously, coordinators would indicate whether the patient met or did not meet each criterion (or if data were insufficient to determine), and if results were discordant, to provide a rationale why the AI was incorrect. Teams used either a junior-to-senior re-assessment when AI–human discordance was detected or single-reviewer adjudication if the original reviewer was senior, according to the trial team’s preference. After the criteria were reviewed, the study staff categorized the patient’s study status (Prescreen Success/Failure, Screening Success/Failure, Patient/Physician Denied, Enrolled); categorization could be later defined if further documentation was needed or if the coordinator was uncertain. Reminder notifications were generated if patients were not reviewed within four days of AI prescreening. If patients were identified outside this workflow, i.e., if a provider needed to be added to the screening list or the Tier 1 AI logic was incorrect, then this prompted either systematic updates to the workflows or adjustments to the AI logic, respectively.

During the first month of a trial utilizing this pipeline, the AI team met with study staff every two weeks to review errors or issues; thereafter, teams met monthly. We implemented a human-in-the-loop, semi-automated AI improvement mechanism that leveraged comments on high-impact issues raised during scheduled meetings and the coordinator’s feedback on the AI’s logic. These improvements in AI logic (ie, criteria prompts) were not reapplied to existing errors. Systemic errors were updated manually, while criteria prompts were updated using a refinement bot (Supplement C: Human-in-the-loop, automated prompt refinement).

### Evaluation and resource utilization

Criterion-level sensitivity, specificity, positive predictive value (PPV), negative predictive value (NPV), and F1-score were computed using human labels as ground truth and tracked over time. Criterion labeled *uncertain* or *no documentation found* by either AI or the coordinators were excluded at the time of analysis. We defined the positive class as {*met, likely met*} and the negative class as {*not met, likely not met*} . False-positive and false-negative cases underwent secondary review within the respective trial team as previously discussed. We report both weighted (by metric-appropriate denominators) and distributional (median, IQR) summaries; 95% confidence intervals for weighted metrics were estimated by bootstrap (1,000 replicates).

We prespecified patient-level outcomes by AI-labeled thresholds (percentage of trial-specific criteria labeled *met* or *likely met* over all criteria): 60–80% and ≥ 80%. For each trial and threshold bin, we calculated the number of prescreen successes, prescreen failures, and unknowns (UNKs). Prescreen success meant the patient successfully passed initial prescreening criteria (based on AI/reviewer); prescreen failure meant the patient did not meet prescreen criteria (based on AI/reviewer). When a patient specific trial label (prescreen outcome) was not explicitly recorded by staff, we applied a deterministic imputation rule using criterion-level coordinator reviews available at the time of analysis: imputed prescreen success if every all criteria were labeled *met* by the coordinator; imputed prescreen failure if any reviewed criterion was labeled *not met* by the coordinator; UNK meant there was a partial review that was incomplete (e.g., remaining criteria *uncertain* or *no documentation found* or *pending review*, which represented a NULL value, ie the coordinator has not placed an evaluation yet for that criterion).

To quantify resource use, we aggregated Azure usage by resource group and normalized to $/patient at the prescreen evaluation level–the cost of running the AI. There were three resource groups covering the cost of this service. We report overall and per-group values (**Table 3**).

## Results

From 10/2024 to 9/2025, a total of 39,182 patients were prescreend by AI, with 914 patients evaluated by trial coordinators in 26 clinical studies. Twenty-one were oncology trials (medical oncology, radiation oncology, neuro-oncology) and five were non-oncologic trials (neurosurgery, mineral and metabolism, hepatology, psychiatry, pulmonology). Further trial characteristics can be found in **Table S2**. AI prescreening was utilized for these studies over a median of 4 months (range: 1-12 months; IQR 3-7 months). Across these trials, 5,096 criteria (112 distinct) were reviewed by trial coordinators (3,880 true positives, 942 true negatives, 220 false positives, and 74 false negatives **Table 1**).

**Table 1.** Overall AI Performance Metrics Across Selected Trials. Analysis includes 112 criteria from 26 trials (5,096 total criteria). TP=3,880, TN=942, FP=220, FN=74. Weighted averages are calculated using sample-size weights with bootstrap confidence intervals (95% CI). Distribution summaries are presented as median and interquartile range (IQR) across criteria (unweighted). *Abbreviations:* TN, True Negative; TP, True Positive; FN, False Negative; FP, False Positive.

| Metric | Weighted Average (95% CI) | Median (IQR) |
| --- | --- | --- |
| Accuracy | 0.942 (0.920–0.960) | 0.984 (0.902–1.000) |
| Sensitivity | 0.981 (0.973–0.988) | 1.000 (0.983–1.000) |
| Specificity | 0.811 (0.709–0.875) | 0.922 (0.625–1.000) |
| Positive Predictive Value | 0.946 (0.919–0.967) | 1.000 (0.931–1.000) |
| Negative Predictive Value | 0.927 (0.896–0.951) | 1.000 (0.833–1.000) |
| F1 Score | 0.966 (0.953–0.977) | 0.991 (0.941–1.000) |

Weighted performance metrics demonstrated high overall accuracy (0.94, 95% CI: 0.92–0.96) and sensitivity (0.98, 95% CI: 0.97–0.99). Specificity was 0.81 (95% CI: 0.71–0.88). PPV and NPV were 0.95 (95% CI: 0.92–0.97) and 0.93 (95% CI: 0.90–0.95), respectively. The overall weighted F1 score was 0.97 (95% CI: 0.95–0.98).

Median metrics across individual criteria indicated similarly high values, with median accuracy of 0.98 (IQR: 0.90–1.00), sensitivity of 1.00 (IQR: 0.98–1.00), and specificity of 0.92 (IQR: 0.63– 1.00). PPV and NPV had medians of 1.00 with IQRs of 0.93–1.00 and 0.83–1.00, respectively. The median F1 score was 0.99 (IQR: 0.94–1.00). Detailed results of aggregate AI performance metrics are summarized in **Table 1. Table S5** in Supplement A: Representative per-criterion performance contains illustrative examples of AI’s ability to address various criteria.

**Table S3** in Supplement A: Per trial screening outcomes summarizes, for each trial, how many patients the AI pathway flagged as potentially eligible under two AI based eligibility thresholds (≥ 80% and 60–80%), the proportion of flagged patients who were reviewed by study staff, and the outcome of that manual prescreening. Across 26 trials, manual review was recorded in our system in 24 (92%) studies. The Leptomeningeal Metastases Prospective Study closed within one month of undergoing AI-based prescreening, and the IDH1-Mutant Recurrent/Progressive Glioma Trial was only recently added at the time of analyses. Patients were more likely to be reviewed by coordinators (544/987, 55.1% vs 372/397, 93.7%) and more likely to be labeled as a prescreen success (104/544, 19.1% vs 162/372, 43.5%) when ≥ 80% criteria were labeled as *met* or *likely met* by AI. Further information can be found in **Table 2**.

**Table 2:** Pooled prescreening outcomes by percentage of criteria labeled *met* or *likely met* by AI across the 24 trials with any coordinator recorded review. *Abbreviations*: HR, Human Reviewed; PS, Prescreen Success; PF, Prescreen Failure; UNK, Unknown.

|  | Bin | Total (N) | HR (N) | PS (N) | PF (N) | UNK (N) | HR / Total (%) | PS / HR (%) | PF / HR (%) | UNK / HR (%) |
| --- | --- | --- | --- | --- | --- | --- | --- | --- | --- | --- |
| AI labeled positive | 60–80% | 985 | 542 | 102 | 401 | 39 | 55.0% | 18.8% | 74.0% | 7.2% |
| AI labeled positive | >80% | 397 | 372 | 162 | 183 | 27 | 93.7% | 43.5% | 49.2% | 7.3% |

**Notes.** HR / Total = proportion reviewed among those flagged by the AI in the bin. PS / HR, PF / HR, and UNK / HR use the number reviewed (HR) as the denominator. Percentages are rounded to one decimal place.

**Table 3.** Azure compute costs for the AI prescreening service by resource group (Payment Groups 1–3). Patients (n) is the number of completed AI prescreen evaluations used to normalize costs for that row. “Total cost” is the Azure invoiced amount (USD). “Cost per patient” equals Total cost*÷* Patients for the same row (USD). The OVERALL row aggregates across all resource groups. Values rounded to two decimals.

| Azure Resource # | Patients, n | Total cost, USD | Cost per patient, USD |
| --- | --- | --- | --- |
| Payment Group 1 | 2,173.00 | 266.32 | 0.12 |
| Payment Group 2 | 6,104.00 | 616.24 | 0.10 |
| Payment Group 3 | 2,319.00 | 400.92 | 0.17 |
| **OVERALL** | 10,596.00 | 1,283.48 | 0.12 |

Since implementing systematic tracking of prompt updates via our semi-automated workflow in April 2025, 27 criteria have been updated at least once based on feedback from trial coordinators. An illustrative example of performance increase over time can be seen in **Figure 1**. In this example from the Newly Diagnosed GBM Adaptive Therapy Trial, the AI’s performance was initially poor for all metrics associated with a criterion that required patients to have *no prior chemotherapy, radiation, or other anti-cancer therapy*. Several early patients who were prescreened were already on trials with unique study drugs (drugs not standard in the treatment of glioblastoma). After implementing our self learning pipeline, the AI learned to be aware that patients could be on trial for their glioblastoma and have non-standard treatments that would make them ineligible. Notable improvement in evaluation metrics was seen from 5/2025 to 8/2025 after this automated learning was implemented in 5/2025 and 6/2025.

**Fig. 1.**
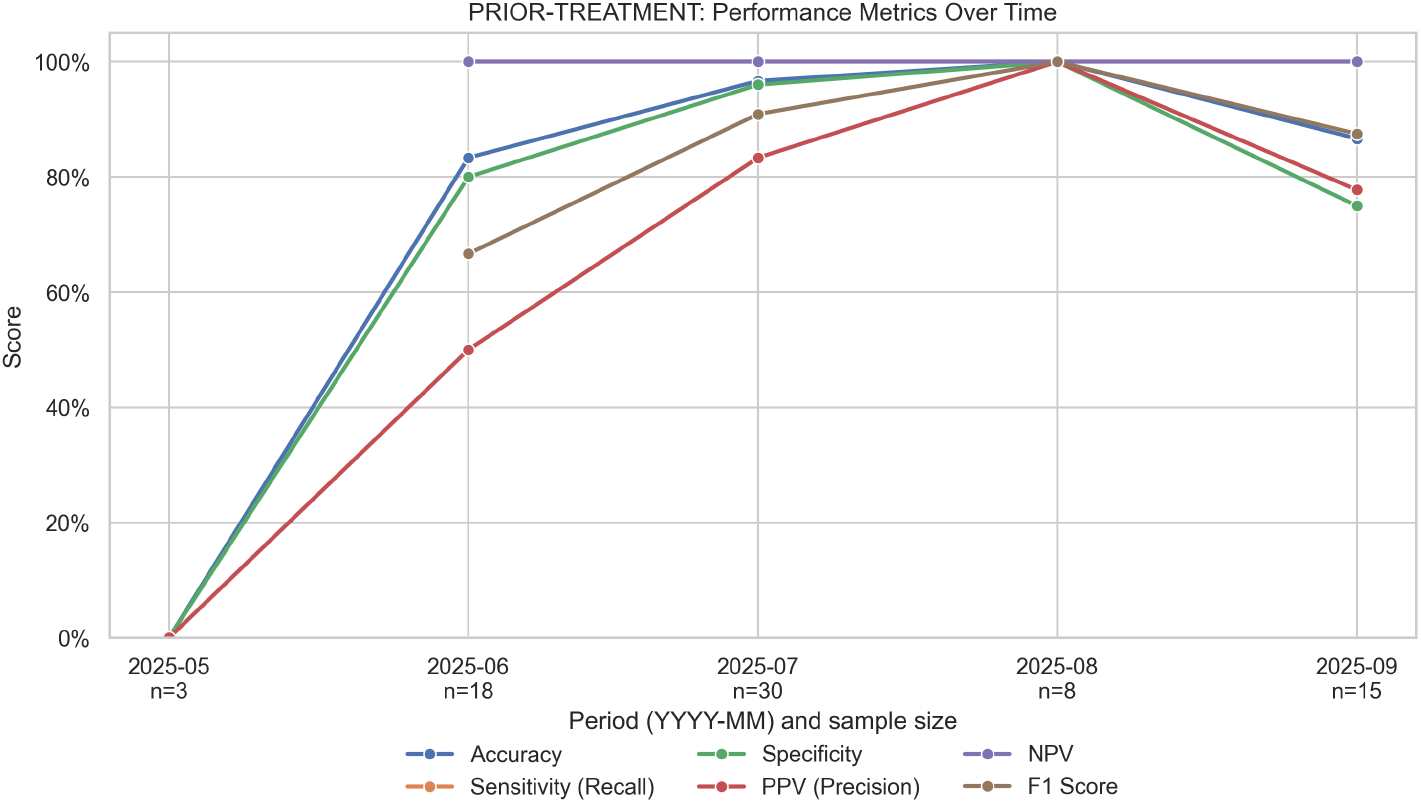
Improved performance over time for a criterion in a primary brain tumor study that required that patients could have *no prior chemotherapy, radiation, or other anti-cancer therapy*. The plot displays monthly accuracy, sensitivity, specificity, PPV, NPV, and F1-score. Early in deployment (May–June 2025), the system under-recognized prior therapy when documented as less common or trial-specific regimens (i.e., beyond standard agents such as temozolomide), leading to false eligibility calls. Following reviewer feedback, retrieval prompts and synonym dictionaries were expanded, and the decision rules were clarified. As a result, all metrics rose steadily and approached ceiling performance by August–September 2025. *PRIOR-TREATMENT* was the short name provided within the coordinator-facing interface. *Abbreviations:* PPV, positive predictive value; NPV, negative predictive value.

**Table 3** shows that the cost was modest: across 10,596 patients prescreen prescreened by AI (sampled from 8/16/2025-9/16/2025), Azure costs totaled $1,283.48, or $0.12 per patient overall (range $0.10–$0.17 across resource groups).

## Discussion

In a prospective deployment across 26 actively enrolling trials spanning oncology and non-oncology domains, we found that a LLM/AI-assisted prescreening workflow is feasible at scale and achieves high agreement with human review at the criterion level. In addition, using LLMs to pre-screen patients identifies subsets of patients most likely to be eligible for clinical trials, increasing the yield of manual coordinator pre-screening efforts. Our initial experience with using this pipeline highlights some potential best practices for other institutions seeking to utilize LLMs for trial prescreening and reveals ongoing difficulties in recruiting patients for clinical trials.

Prior studies have focused either on accuracy in controlled settings or on single-trial deployments[10–15]. For example, *TrialGPT* reported criterion-level accuracy of 87.3% in manual evaluations of 1,015 synthetically generated patient–criterion pairs[10]. In heart failure, *RECTIFIER* was tested against blinded clinician standards and study staff: question-level accuracy for RECTIFIER ranged 97.9–100% with overall sensitivity/specificity of 92.3%/93.9%[11] with very low per-patient costs (about $0.11 for a single-question strategy). A subsequent randomized comparison within the same program reported faster eligibility determinations and more enrollments with AI assistance[12]. In oncology, *OncoLLM* reported 75.3% accuracy in a 720-item criterion set derived from 10,000 notes/50 patients[15]. Taken together, these studies demonstrate promise but also illustrate that accuracy depends on data, prompts, model choice, and evaluation design. Our deployment—across 26 trials and 112 distinct criteria—showed high aggregate agreement with human review at the criterion level (accuracy *>*94%). We did not have AI make a *global* patient-level eligibility, as accuracy typically declines when multiple criteria must be satisfied simultaneously. Our findings should therefore be interpreted as criterion-level performance within routine screening, not as patient-level eligibility determinations.

Recognizing that no LLM screen is 100% accurate, our institution required that all AI-flagged candidates undergo coordinator review before the trial-specific research team reached out to providers regarding a patient’s potential eligibility. To balance sensitivity with coordinator effort, study teams used a variable review threshold based on the proportion of trial-specific criteria labeled *met* or *likely met* by AI; in practice, teams more commonly reviewed patients at or above 50-60% and prioritized those above 75-80% even though no limitations were placed to review patients below 50%. These thresholds were intended to route charts to human review rather than to make enrollment decisions. Consistent with that intent, we observed higher rates of completed reviews among patients with an AI labeled threshold of ≥ 80%. One significant limitation to this approach is if AI does not label the initial main criterion (Tier 1) as *met* or *likely met* then the other criteria will not be evaluated and by design they will not meet even the 50% threshold, making them less likely to be reviewed. This poses an issue if the AI was incorrect and therefore there is likely an under-representation of false negatives. We asked coordinators if patients were identified outside of this workflow; if so, we would review for false negatives and apply our human-in-the-loop learning approach.

Free-text notes carry much of the clinical story; however, several criteria in our deployment showed that some signals (for example, active medications and the most recent laboratory values) are captured more reliably in structured EHR tables at the time of evaluation, as opposed to our solely RAG-based approach. A practical extension is an agentic controller—an AI-based helper that chooses where to look first for each criterion—rather than sending all questions through the same retrieval pathway. For criteria that map cleanly to structured data (for example, *currently on SSRI/SNRI* or *latest testosterone above threshold* ), the controller would check the relevant list or lab table first and use notes only to add context. For narrative criteria that depend on clinical reasoning in the chart, it would use retrieval-augmented review of notes. For mixed criteria, it would pull the structured part directly and reserve the note review for the narrative component. Timing was a recurring source of error. Notes often contain older statements that look relevant even when newer information elsewhere tells a different story. To mitigate this, we employed methods to sort documents by relevance and time (node post-processing); however, these errors persisted. For truly time-sensitive checks—such as *latest testosterone* or *currently prescribed antidepressant* —a quick, direct look at the structured lab results or medication list is more reliable, with note review used to explain or confirm the finding. In that sense, timing problems are partly a limitation of text search unless recency is handled explicitly.

Finally, some questions are hard for both humans and AI and require a conscious trade-off. In the escitalopram–asthma study, differentiating an asthma exacerbation from decompensation due to COPD or another illness in a patient with a history of prior asthma exacerbations often required clinician input. In this instance, we favored sensitivity over specificity to avoid missed opportunities; however, decisions like this should be made on a per-trial basis.

Introducing AI into trial screening is as much a people-and-process change as it is a technical one. In our deployment, the AI pre-filter raised the share of coordinator-reviewed charts that were plausibly eligible while quietly setting aside large numbers of likely ineligible patients. The system ran several times each week and aimed to alleviate coordinators’ work; the average computing cost was approximately $0.12 per patient. Teams arrived with different habits—some had already received targeted referrals from engaged clinicians, while others reviewed all patients scheduled with certain providers. Generally speaking, all teams thought an AI-supported workflow was beneficial. It took time to learn how to effectively convey the importance of reviewer feedback to AI; however, once we demonstrated that the workflow could learn and improve, people became more willing to provide that feedback.

### Limitations

First, our analyses were descriptive and not stratified to test whether documentation patterns or local review practices modified outcomes; we therefore restrict claims to feasibility, high criterion-level agreement, and within-trial associations with AI confidence. Second, the tiered criteria processing design used a hard gate at Tier 1: if Tier 1 was labeled anything other than *met* or *likely met*, downstream criteria were not evaluated for that run, and the case appeared lower on the reviewer worklist. This conserves reviewer time but makes Tier 2 assessment conditional on Tier 1 and can reduce opportunities to overturn false negatives. Third, per-criterion performance metrics were calculated only on items with definitive or semi-definitive labels by both AI and human ( *met, likely met, likely not met, not met* ); items labeled *uncertain, need further documentation*, or *no documents found* by either party were excluded. Fourth, scanned external records were excluded by policy and look-back windows were typically 0-180 days, all of which can influence observed prescreen success. Finally, we did not measure effects on approach, consent, enrollment, or accrual. While a randomized comparison in heart failure reported faster eligibility determinations and increased enrollments with AI assistance[12], our data do not address accrual, and future work should include prospective instrumentation of downstream recruitment steps.

## Conclusion

Across 26 trials, an LLM-assisted, human-in-the-loop prescreening workflow proved feasible at scale, achieving greater than 94% criterion-level agreement with coordinator review at approximately $0.12 per patient. AI triage enriched coordinator worklists for plausibly eligible patients while preserving mandatory human oversight and avoiding global patient-level determinations. Structured coordinator feedback enabled automated learning. Future work should measure downstream recruitment outcomes.

## Data Availability

Certain data produced in the present study are available upon reasonable request to the authors

## Acknowledgments

We thank the research coordinators and staff who made this study possible through their diligence in data collection, patient coordination, and study operations. In particular, we are grateful to the following team members (alphabetical): *Alyssa Truxon; Amy Rowell; Aniah Daniels; Andy Gonzalez; Bart Weger; Busayo Adefalujo; Christian Chukwuma; Cristian Gonzalez; Deniz Durakoglugil; Emerald Pina; Erica Salcedo; Guruveer Bhamra; Hannah Pitts; Jonathan VanPelt; Julia Terrell; Kuntal Majmudar; Lisa Quirk; Lily Robles; Lindsay Apgar; Maggie Stein; Noah Puente; Reagan Volzer; Sarah Neufeld; Sneha Deodhar; Tash Mupambo; Tammy Ricklefs*. We are grateful for their contributions to recruitment, monitoring, and day-to-day study support.

## Supplement A

**Table S1:** Example Inclusion / Exclusion Criteria and Example Output from AI Pipeline.

| Inclusion Criteria | Example Output |
| --- | --- |
| Patient must have a CT scan showing either unilateral convexity chronic subdural hematoma (CSDH) measuring at least 10 mm in thickness or bilateral CSDH if only one side is considered for treatment and the contralateral side is asymptomatic and < 5 mm in thickness. | Status: Met<br>Certainty: Very High<br>Support: The overall criterion is met based on clear, direct, and consistent evidence from multiple imaging reports on 07/01/2025 showing a unilateral left frontoparietal extra-axial collection measuring 11 mm in thickness, consistent with a chronic subdural hematoma. The collection causes mild local mass effect but no midline shift or hydrocephalus. There is no evidence of bilateral subdural hematomas or contralateral collections. Clinical history supports the diagnosis. The documentation is extensive, consistent, and unambiguous, providing very high certainty that the criterion is fulfilled. |
| $\geq 3$ asthma exacerbations in the past year requiring $\geq 3$ days of systemic corticosteroids each. | Status: Not Met<br>Certainty: Very High<br>Support: The patient has a diagnosis of asthma and has had some ER visits for asthma exacerbations, but the documentation clearly states only a couple of ER visits and no hospital admissions in the past year. There is no explicit or implicit evidence of systemic corticosteroid treatment for these exacerbations, either in medication lists or progress notes. No systemic corticosteroid prescriptions or courses were documented. The absence of $\geq 3$ exacerbations and absence of systemic corticosteroid treatment for $\geq 3$ days per exacerbation strongly supports that the criterion is not met. This conclusion aligns with the trial criteria and the provided tips that require explicit or inferred evidence of $\geq 3$ exacerbations treated with systemic corticosteroids. Therefore, the criterion is definitively not met with very high certainty. |
| Patient must have deleterious homologous recombination repair (HRR) alteration(s) per any validated test, next generation sequencing (NGS) mutational analysis (tissue or liquid). These include BRCA1/2, BRIP1, CHEK2, FANCA, PALB2, RAD51B, and/or RAD54L. | Status: No Documents Found<br>Certainty: Low<br>Support: The criterion requires documented deleterious HRR alterations in specific genes confirmed by validated tests such as NGS. The patient's records include detailed oncologic history, imaging, biopsies, and treatment notes but lack any genetic testing or molecular profiling data relevant to HRR gene alterations. No references to BRCA1/2, BRIP1, CHEK2, FANCA, PALB2, RAD51B, or RAD54L gene status or mutational analysis were identified. Without any genetic or molecular testing documentation, it is impossible to determine if the patient meets or does not meet the criterion. Therefore, the overall status is "no documents found" with low certainty due to absence of relevant data. |
| Clinical stage I–IVB (AJCC, 7th edition); stages I–II glottic cancer are excluded | Status: Met<br>Certainty: Very High<br>Support: The patient has a clinical stage of cT3N1 for oropharyngeal cancer, which aligns with the AJCC 7th edition criteria for stage I–IVB. There is no mention of glottic cancer involvement, specifically stages I–II, in the documents. The evidence is clear and consistent across multiple documents, supporting the criterion being met with very high certainty. |

| Inclusion / Exclusion Criteria | Example Output |
| --- | --- |
| Rectal tumor must occupy less than half of the circumference | Status: Likely Met<br>Certainty: High<br>Support: The overall evidence from multiple imaging reports (MRI Pelvis 10/04/2024, 04/18/2025; CTAP 02/26/2025) and endoscopic findings indicates no discrete mass or large circumferential tumor. Descriptions of “circumferential wall thickening” are likely related to inflammation or fibrosis rather than tumor burden. No explicit quantification of tumor circumferential involvement is documented, but the residual tumor is described as limited and non-mass-forming. No evidence suggests tumor involvement of 50% or more of the rectal circumference. According to the trial criteria and Tip#2, this supports a conclusion that the tumor likely occupies less than half the circumference. The absence of explicit statements leads to a “likely met” status rather than definitive “met.” The certainty is high (4) due to consistent imaging and clinical descriptions across multiple reports without contradictory evidence. |
| Patients must be Child–Pugh score A within one month prior to study entry. | Status: Met<br>Certainty: Very High<br>Support: The criterion requires a Child–Pugh score A (5–6 points) within one month prior to study entry (2025-09-07). No explicit Child–Pugh score is documented. However, recent labs from 09/03/2025 show total bilirubin < 0.3 mg/dL and albumin 3.9 g/dL, both scoring 1 point each. Clinical notes and imaging within the timeframe show no ascites or hepatic encephalopathy, scoring 1 point each. INR/PT values are not documented, creating some uncertainty, but absence of clinical coagulopathy or bleeding suggests likely INR < 1.7 (1 point). Thus, the estimated Child–Pugh score is likely 5 points, consistent with Class A. The labs and clinical assessments are within one month prior to study entry, meeting the timeframe requirement. Given the strong supporting evidence and minimal ambiguity, the overall criterion is MET with high certainty. This assessment aligns with the trial criteria and tips for calculating Child–Pugh score when explicit documentation is absent (Tips sections: Criterion MET if calculated score 5–6 within one month; absence of ascites and encephalopathy; bilirubin and albumin thresholds). Missing INR data is noted but does not outweigh the overall evidence. [Progress Notes_2025-09-02, Lab_09-03-2025, Imaging_06-30-2025, Tips]. |

**Table S2:**
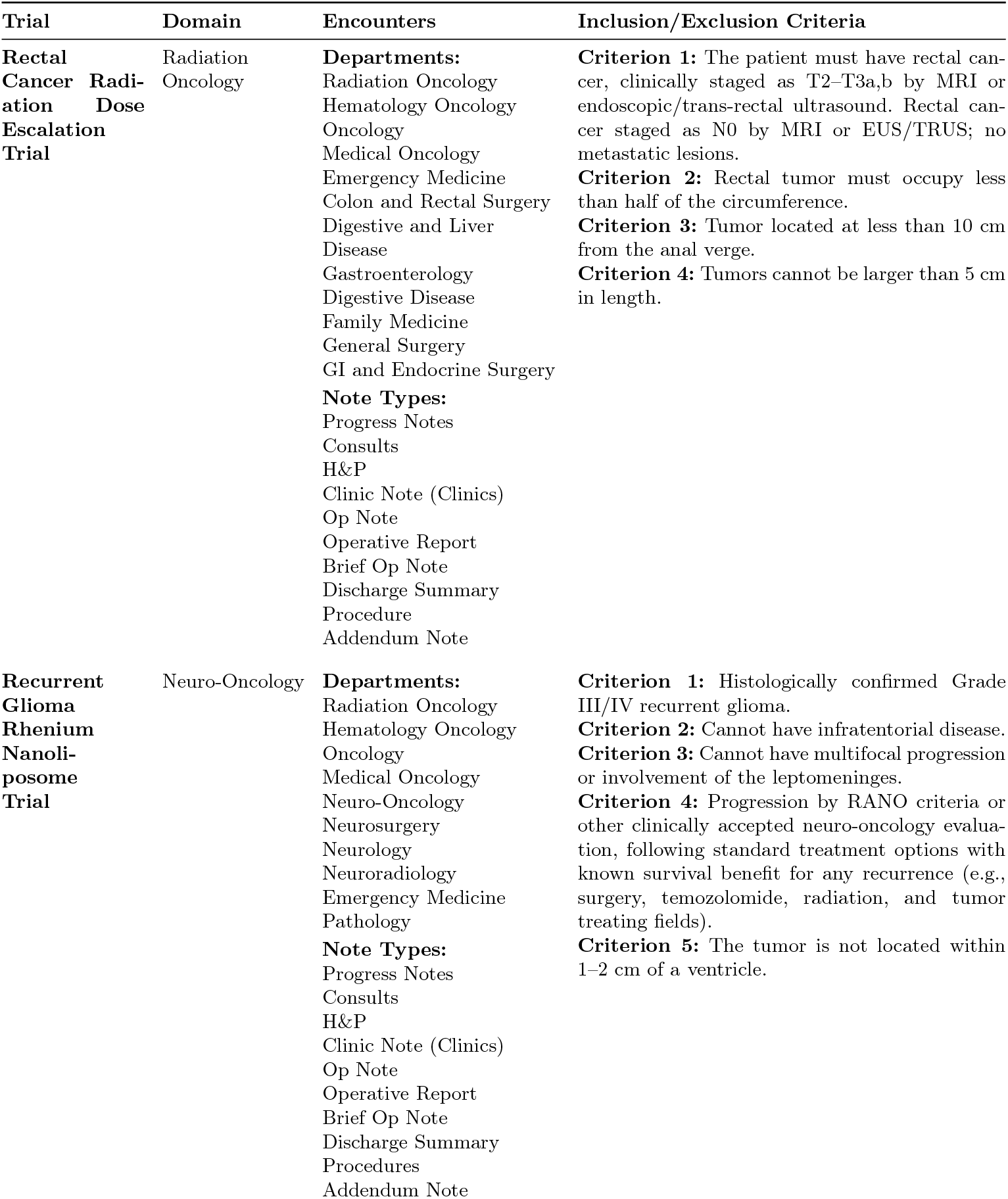
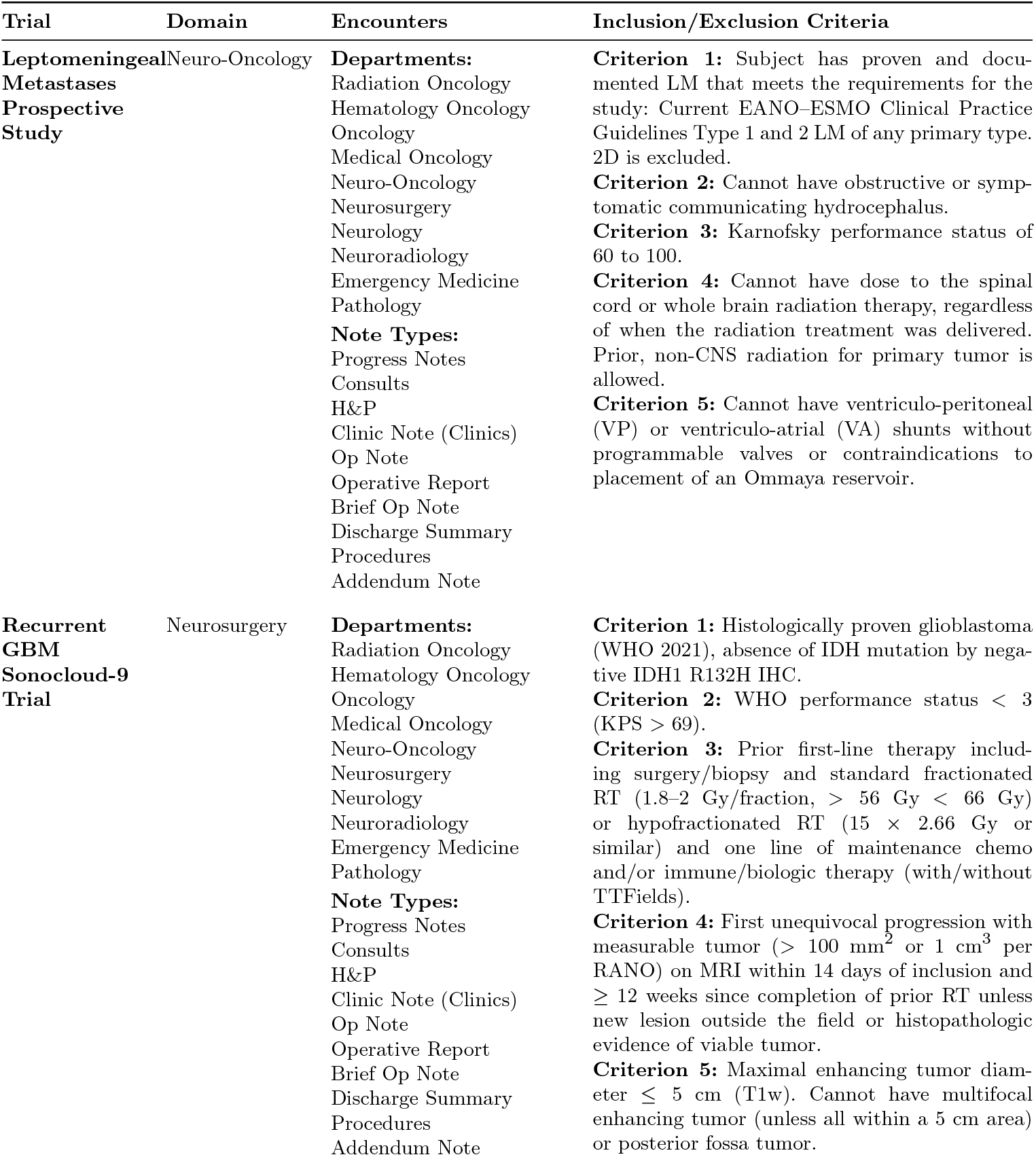
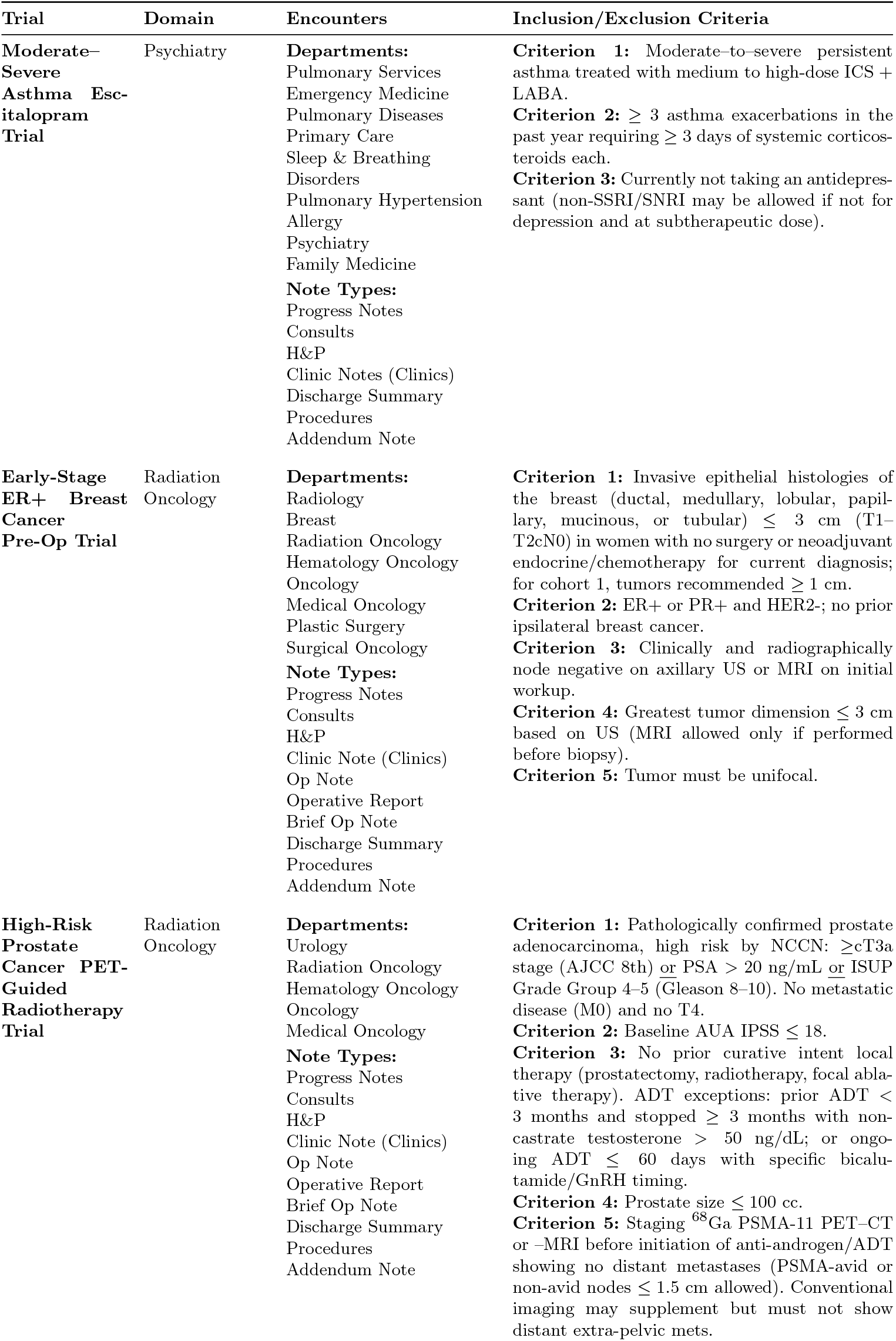
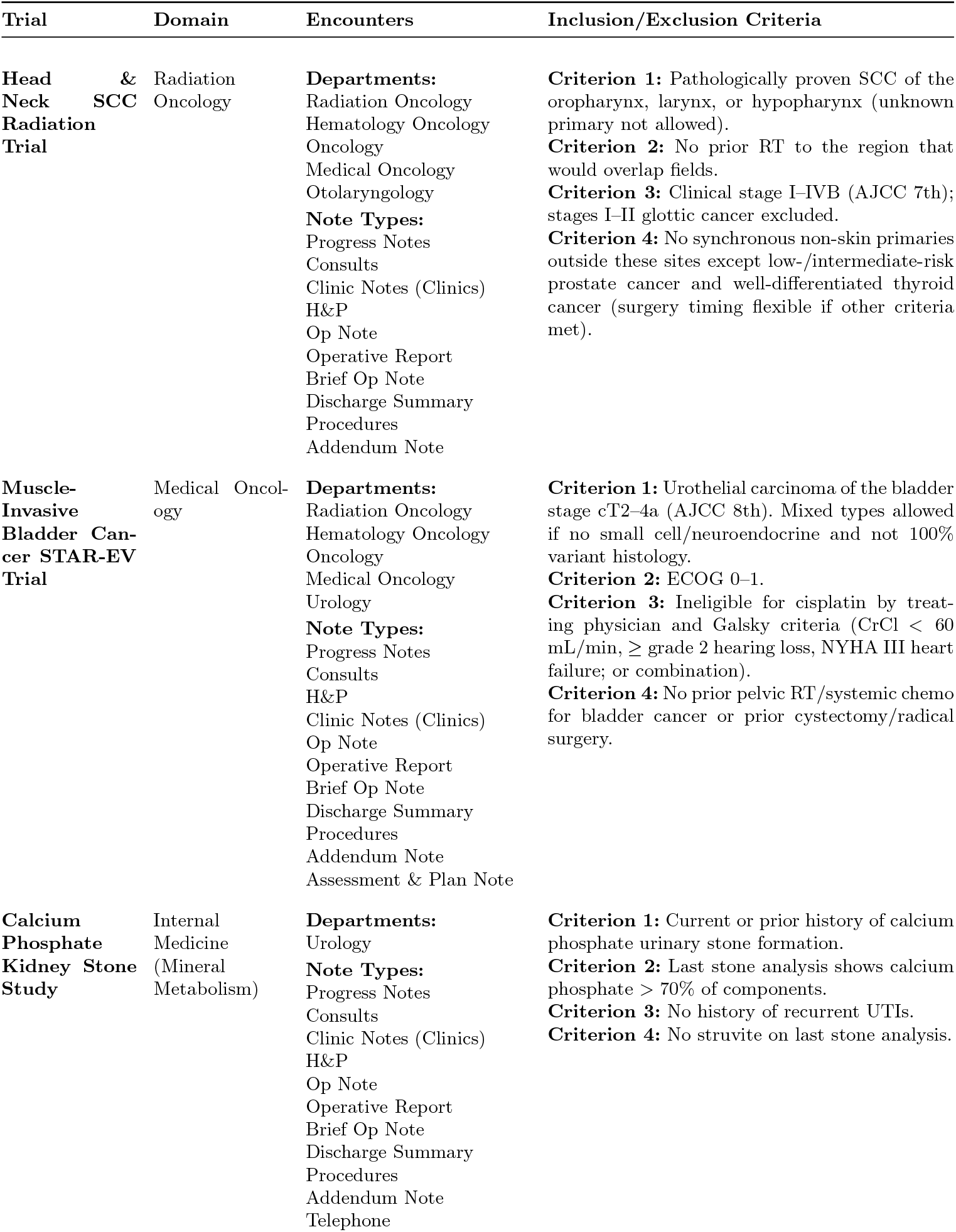
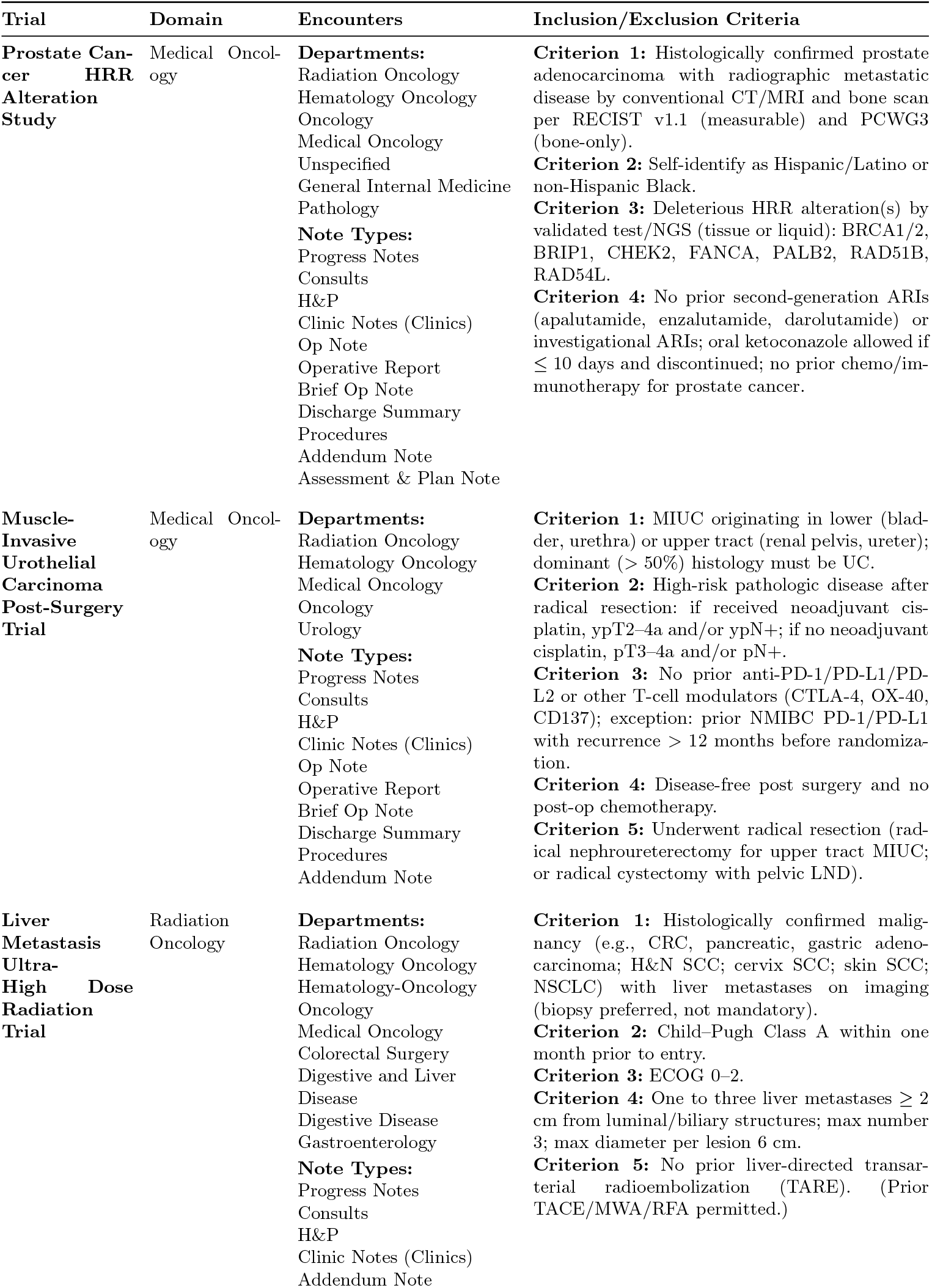
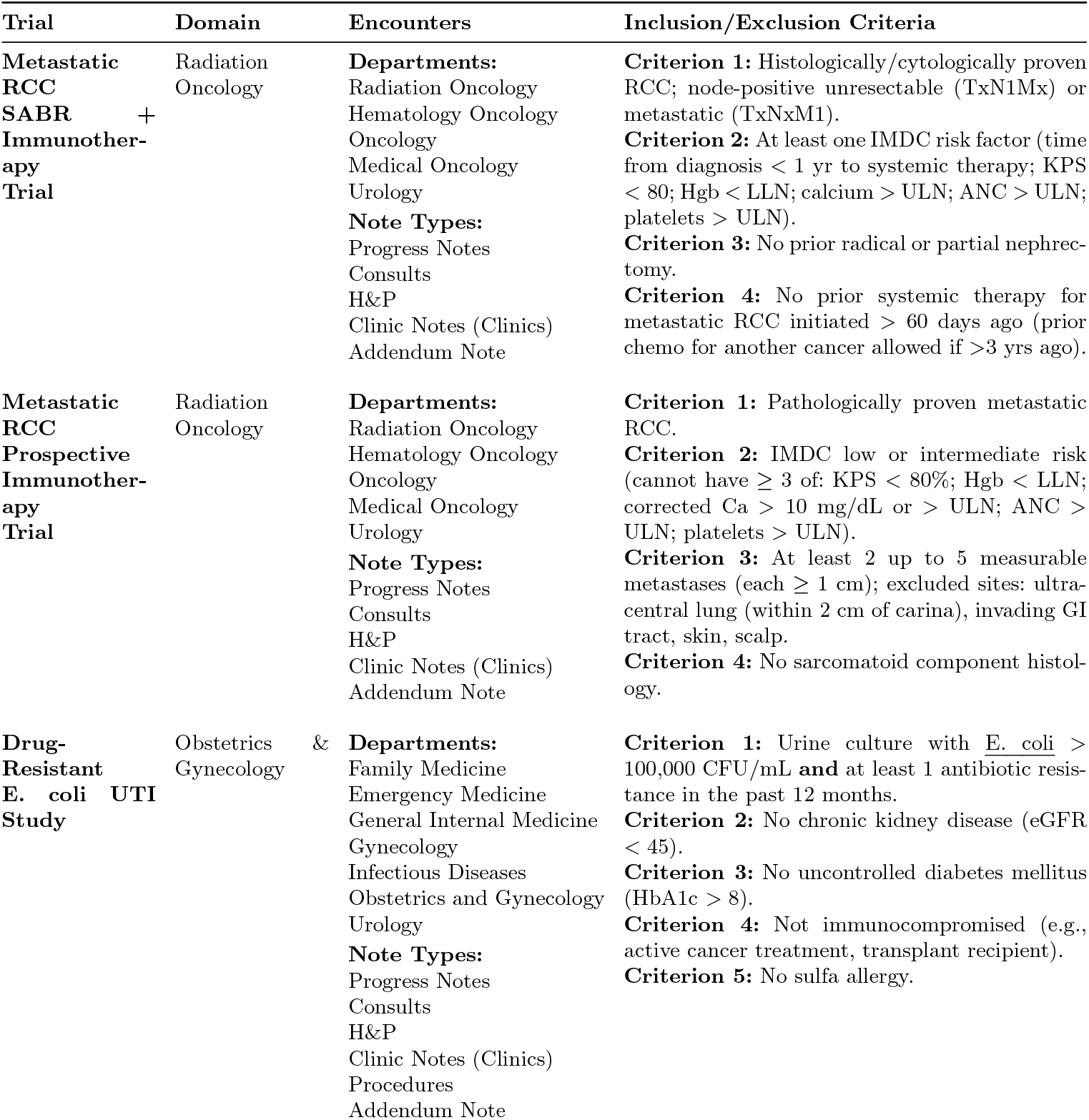
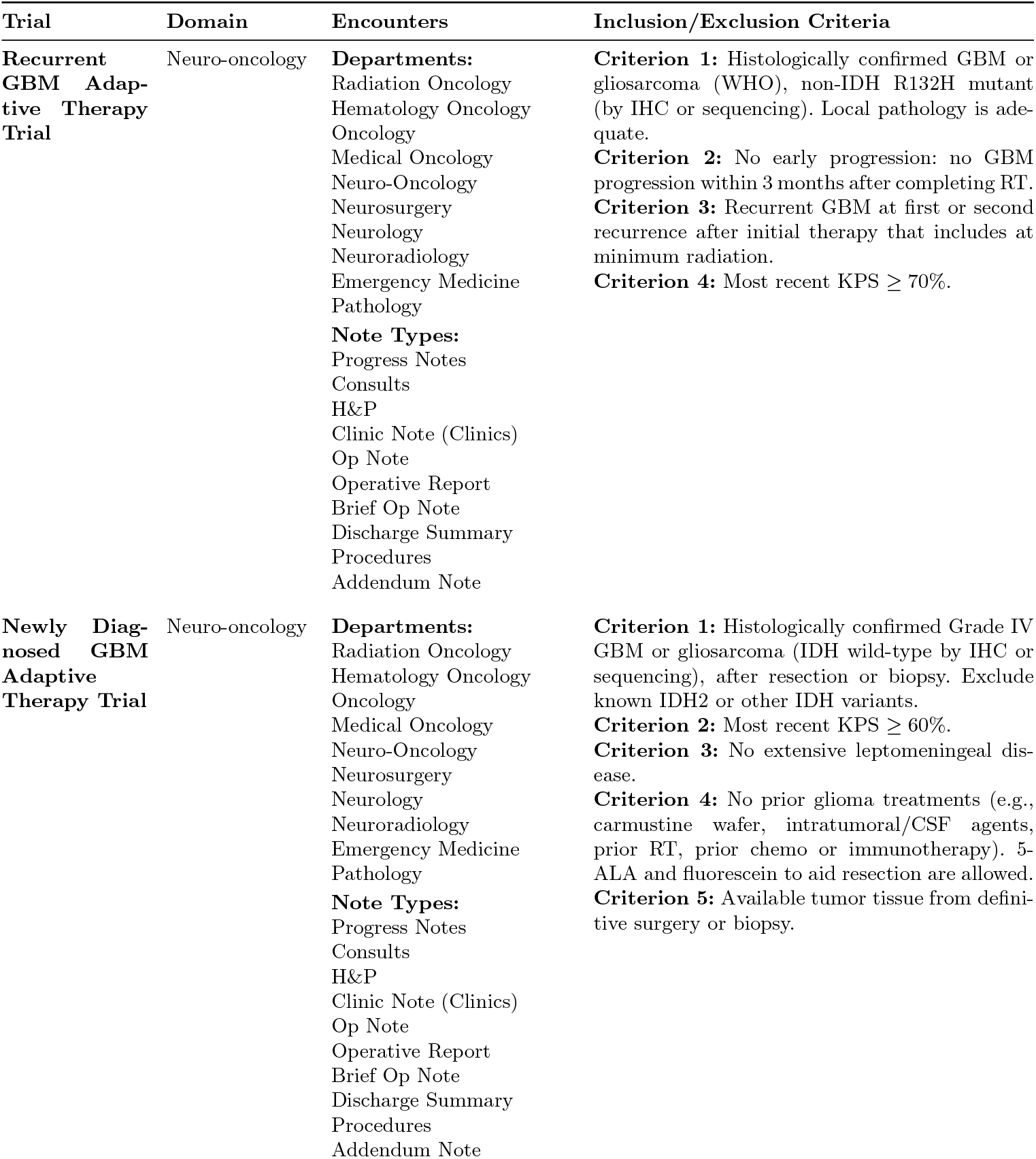
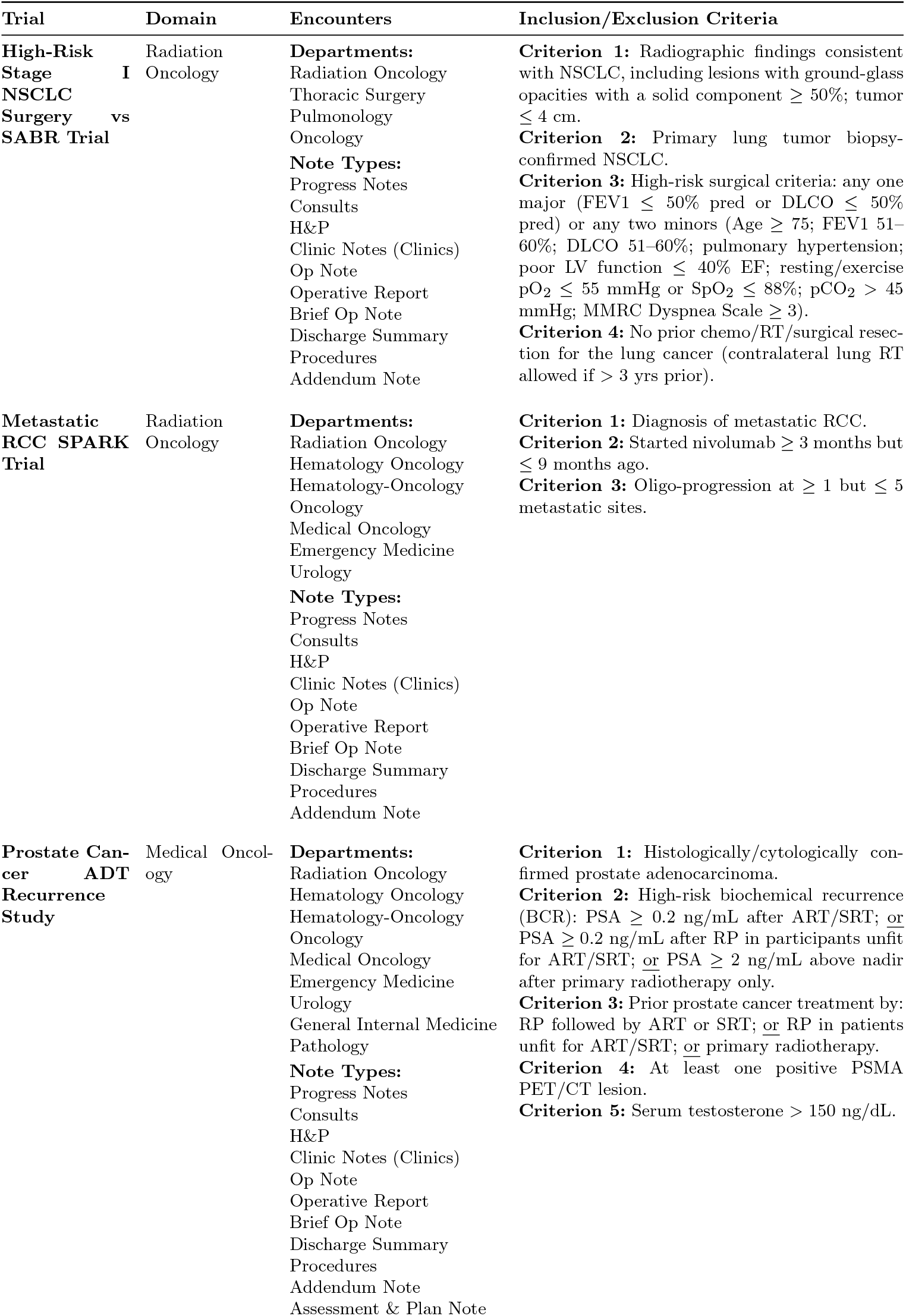
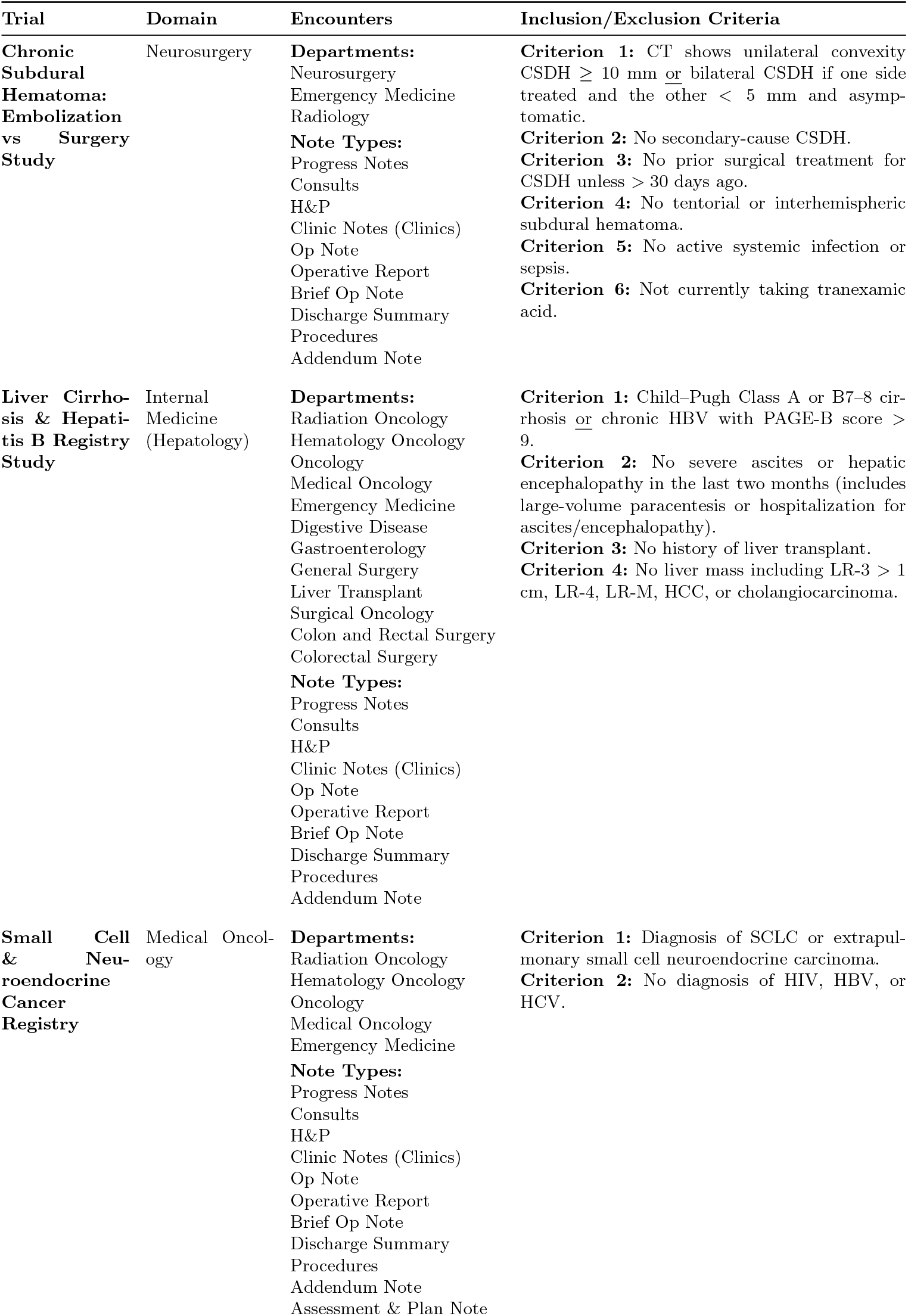
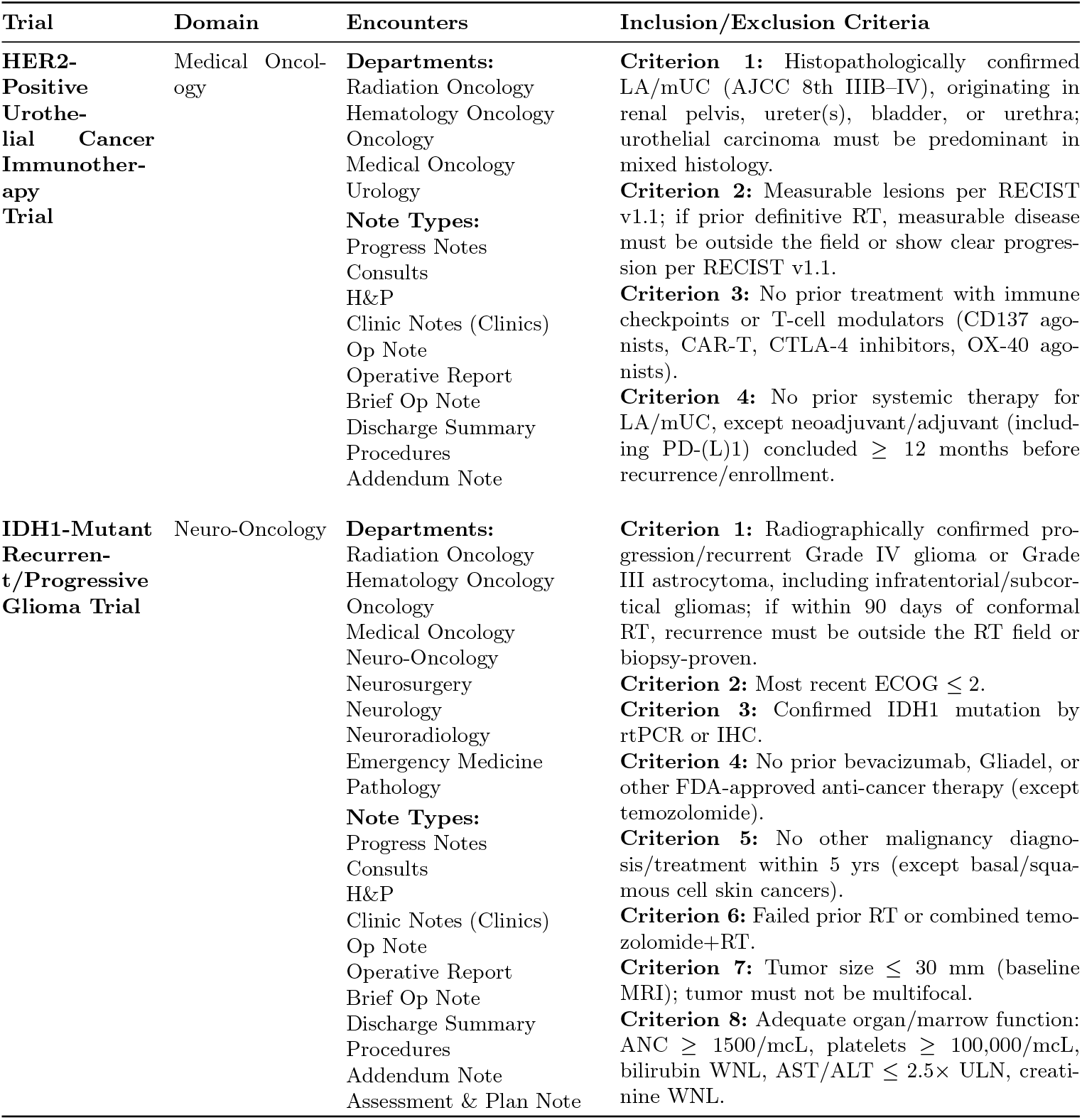
Trials, Domains, Encounters, and Inclusion/Exclusion Criteria. **Abbreviations:** ER+, estrogen receptor–positive; PET, positron emission tomography; SCC, squamous cell carcinoma; GBM, glioblastoma; UTI, urinary tract infection; NSCLC, non–small cell lung cancer; SABR, stereotactic ablative radiotherapy; RCC, renal cell carcinoma; ADT, androgen deprivation therapy; HRR, homologous recombination repair; HER2, human epidermal growth factor receptor 2; IDH1, isocitrate dehydrogenase 1; *E. coli, Escherichia coli* ; Pre-Op, preoperative; vs, versus.

**Table S3:**
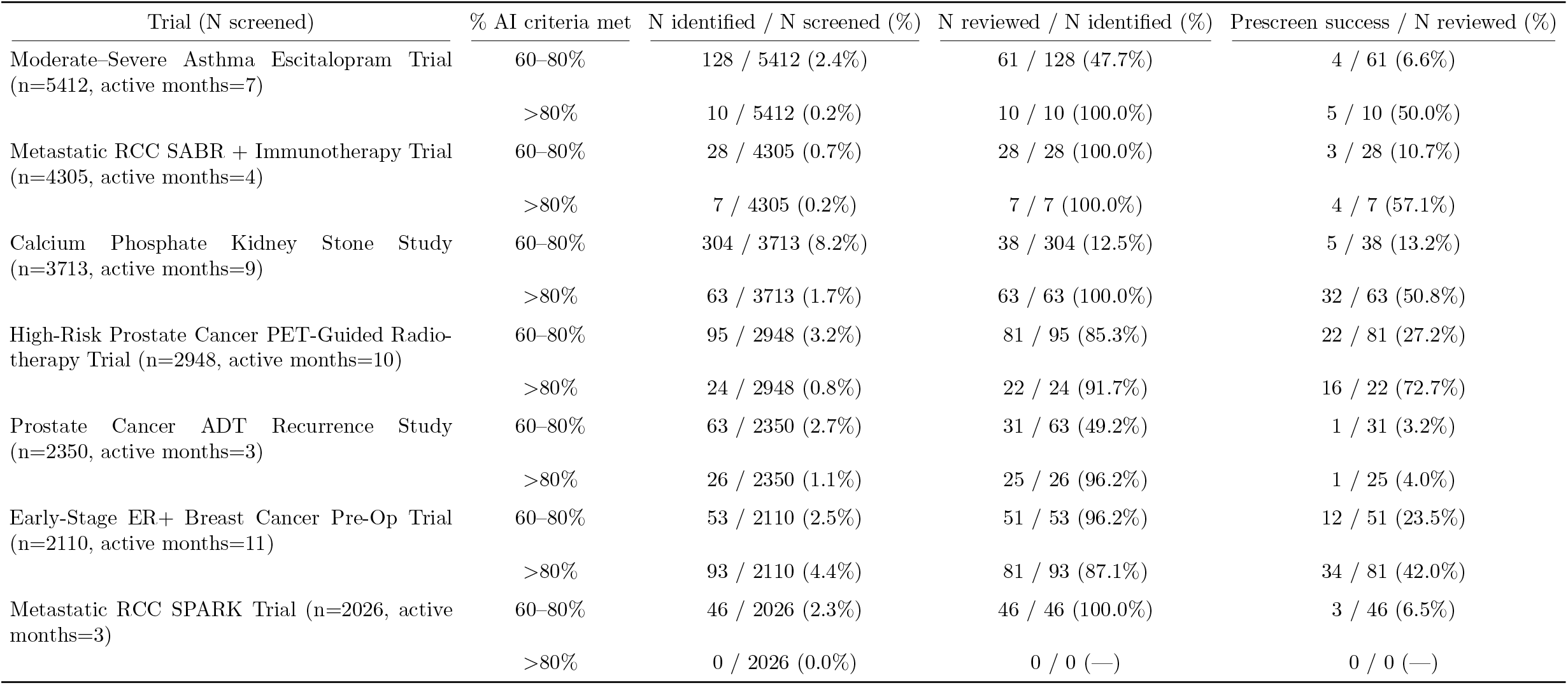
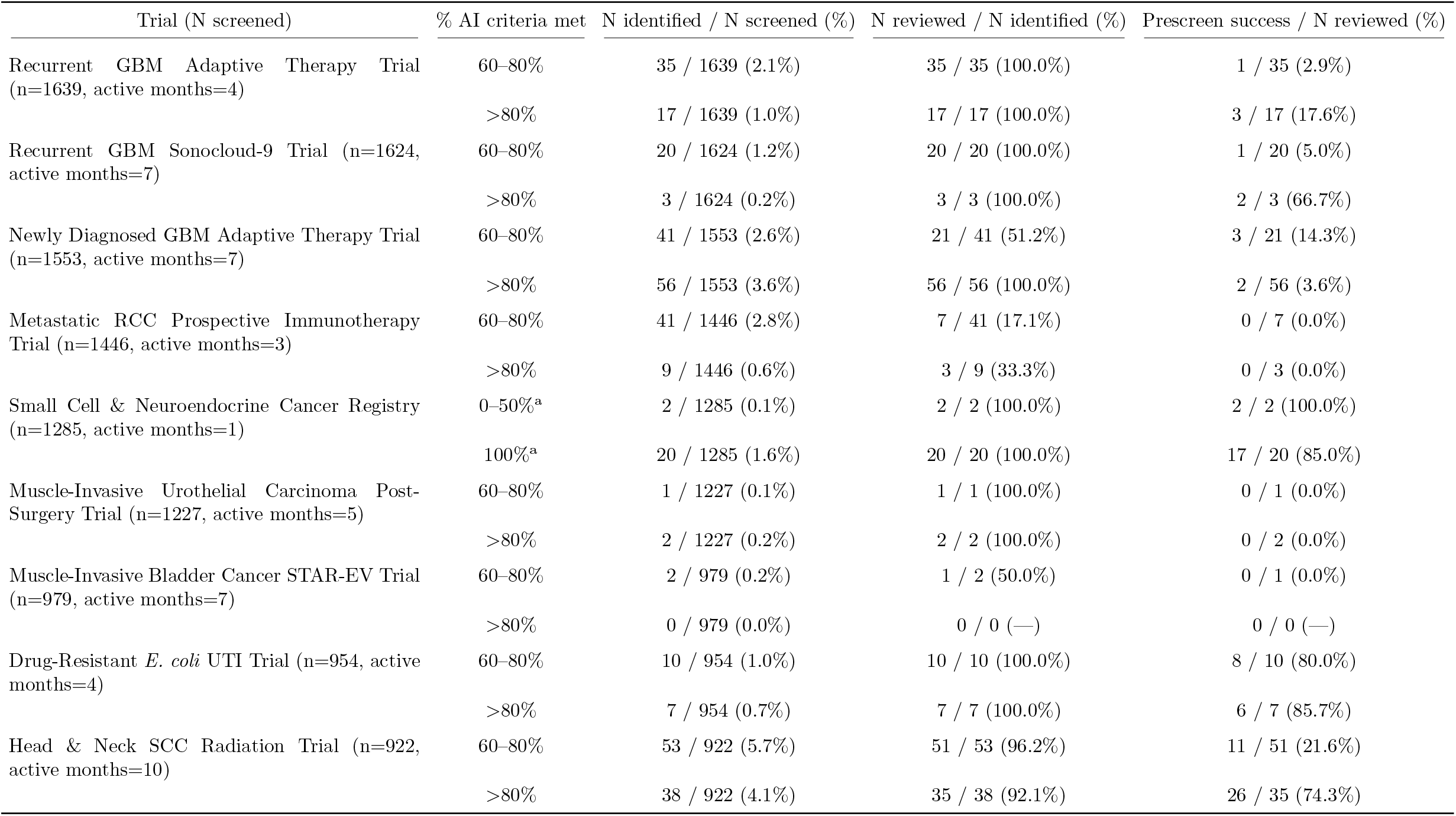
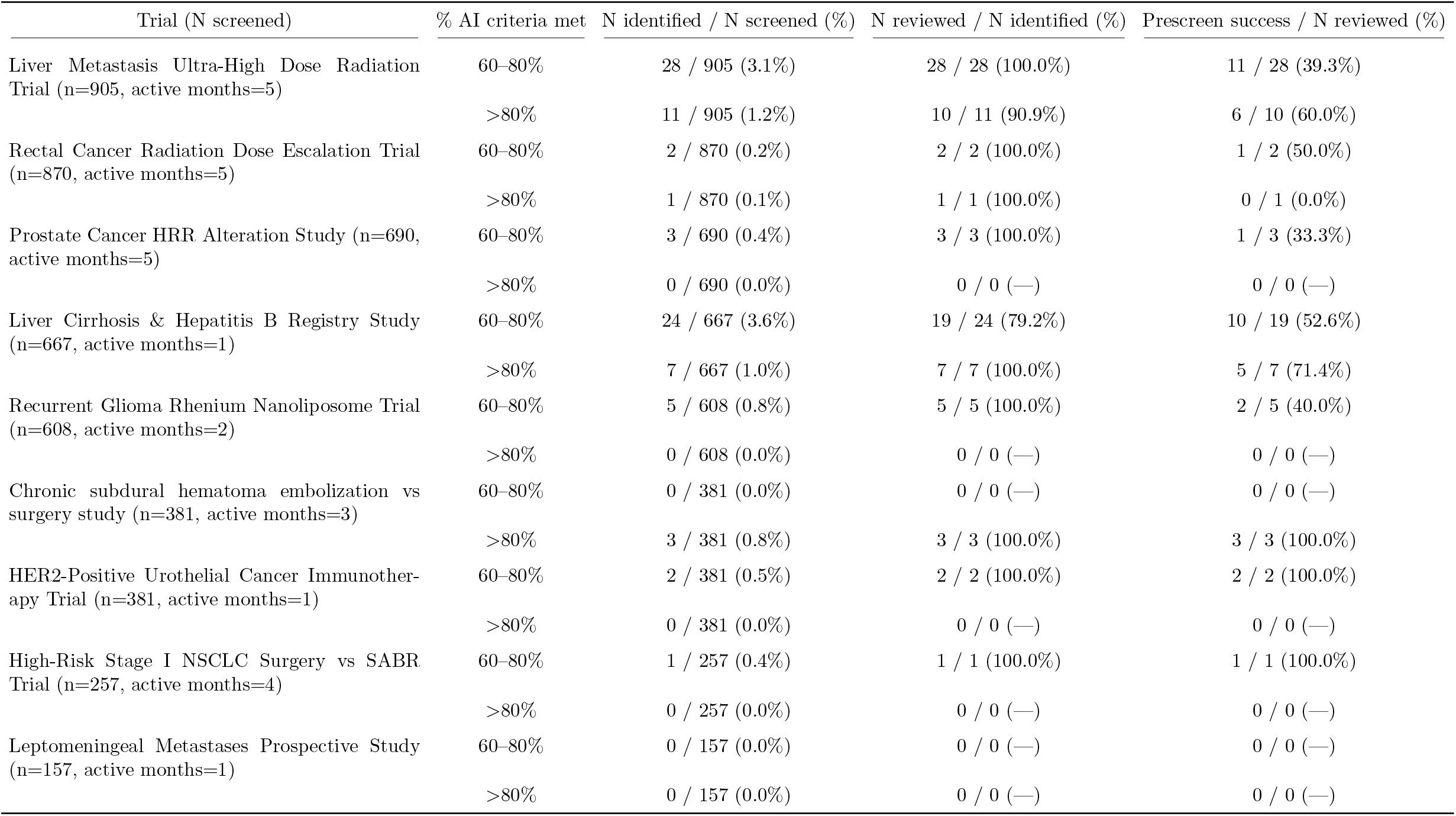
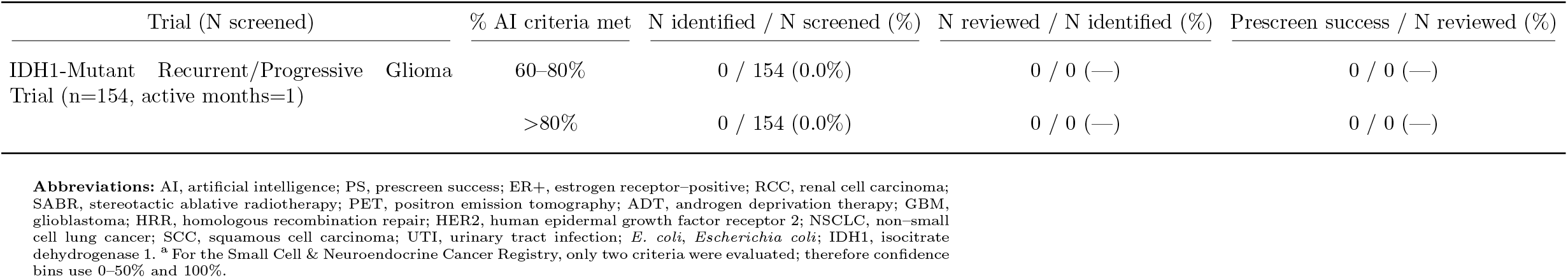
AI-assisted prescreening workflow across trials, reported by AI-confidence bin. For each trial (*n* = patients screened by the AI for that trial), the columns report: (1) N identified / N screened (%) — patients flagged by the AI within the stated confidence bin divided by all patients screened by the AI for that trial; (2) N reviewed / N identified (%) — unique patients (with any human review recorded) among those identified by the AI in the bin; and (3) Prescreen success / N reviewed (%) — patients deemed eligible at the prescreen stage among those reviewed. Rows are ordered by the number of patients screened by the AI for the trial (*n*). Percentages use the denominator shown after the slash in each cell; “—” denotes not applicable (denominator = 0).

**Table S4:** Prescreen failure (PF) and unknown (UNK) outcomes by trial and AI match threshold. PF is reported as Prescreen failure / N reviewed (%); UNK is reported as Unknown / N reviewed (%). Trials are ordered by the number of patients screened by the AI (*n*).

| Trial (N screened) |  |  |  |  | % AI criteria met | Prescreen failure / N reviewed (%) | Unknown / N reviewed (%) |
| --- | --- | --- | --- | --- | --- | --- | --- |
| Moderate–Severe Asthma Escitalopram Trial (n=5412) |  |  |  |  | 60–80% | 57 / 61 (93.4%) | 0 / 61 (0.0%) |
|  |  |  |  |  | >80% | 5 / 10 (50.0%) | 0 / 10 (0.0%) |
| Metastatic RCC SABR + Immunotherapy Trial (n=4305) |  |  |  |  | 60–80% | 25 / 28 (89.3%) | 0 / 28 (0.0%) |
|  |  |  |  |  | >80% | 3 / 7 (42.9%) | 0 / 7 (0.0%) |
| Calcium Phosphate Kidney Stone Study (n=3713) |  |  |  |  | 60–80% | 33 / 38 (86.8%) | 0 / 38 (0.0%) |
|  |  |  |  |  | >80% | 31 / 63 (49.2%) | 0 / 63 (0.0%) |
| High-Risk Prostate Cancer PET-Guided Radiotherapy Trial (n=2948) |  |  |  |  | 60–80% | 59 / 81 (72.8%) | 0 / 81 (0.0%) |
|  |  |  |  |  | >80% | 6 / 22 (27.3%) | 0 / 22 (0.0%) |
| Prostate Cancer ADT Recurrence Study (n=2350) |  |  |  |  | 60–80% | 30 / 31 (96.8%) | 0 / 31 (0.0%) |
|  |  |  |  |  | >80% | 24 / 25 (96.0%) | 0 / 25 (0.0%) |
| Early-Stage ER+ Breast Cancer Pre-Op Trial (n=2110) |  |  |  |  | 60–80% | 39 / 51 (76.5%) | 0 / 51 (0.0%) |
|  |  |  |  |  | >80% | 47 / 81 (58.0%) | 0 / 81 (0.0%) |
| Metastatic RCC SPARK Trial (n=2026) |  |  |  |  | 60–80% | 43 / 46 (93.5%) | 0 / 46 (0.0%) |
|  |  |  |  |  | >80% | 0 / 0 (—) | 0 / 0 (—) |
| Recurrent GBM Adaptive Therapy Trial (n=1639) |  |  |  |  | 60–80% | 5 / 35 (14.3%) | 29 / 35 (82.9%) |
|  |  |  |  |  | >80% | 14 / 17 (82.4%) | 0 / 17 (0.0%) |

| Trial (N screened) | % AI criteria met | Prescreen failure / N reviewed (%) | Unknown / N reviewed (%) |
| --- | --- | --- | --- |
| Recurrent GBM Sonocloud-9 Trial (n=1624) | 60–80% | 10 / 20 (50.0%) | 9 / 20 (45.0%) |
|  | >80% | 0 / 3 (0.0%) | 1 / 3 (33.3%) |
| Newly Diagnosed GBM Adaptive Therapy Trial (n=1553) | 60–80% | 18 / 21 (85.7%) | 0 / 21 (0.0%) |
|  | >80% | 30 / 56 (53.6%) | 24 / 56 (42.9%) |
| Metastatic RCC Prospective Immunotherapy Trial (n=1446) | 60–80% | 7 / 7 (100.0%) | 0 / 7 (0.0%) |
|  | >80% | 3 / 3 (100.0%) | 0 / 3 (0.0%) |
| Small Cell & Neuroendocrine Cancer Registry (n=1285) | 0–50% | 0 / 2 (0.0%) | 0 / 2 (0.0%) |
|  | 100% | 2 / 20 (10.0%) | 1 / 20 (5.0%) |
| Muscle-Invasive Urothelial Carcinoma Post-Surgery Trial (n=1227) | 60–80% | 0 / 1 (0.0%) | 1 / 1 (100.0%) |
|  | >80% | 1 / 2 (50.0%) | 1 / 2 (50.0%) |
| Muscle-Invasive Bladder Cancer STAR-EV Trial (n=979) | 60–80% | 1 / 1 (100.0%) | 0 / 1 (0.0%) |
|  | >80% | 0 / 0 (—) | 0 / 0 (—) |
| Drug-Resistant *E. coli* UTI Trial (n=954) | 60–80% | 2 / 10 (20.0%) | 0 / 10 (0.0%) |
|  | >80% | 1 / 7 (14.3%) | 0 / 7 (0.0%) |
| Head & Neck SCC Radiation Trial (n=922) | 60–80% | 40 / 51 (78.4%) | 0 / 51 (0.0%) |
|  | >80% | 9 / 35 (25.7%) | 0 / 35 (0.0%) |
| Liver Metastasis Ultra-High Dose Radiation Trial (n=905) | 60–80% | 17 / 28 (60.7%) | 0 / 28 (0.0%) |
|  | >80% | 4 / 10 (40.0%) | 0 / 10 (0.0%) |
| Rectal Cancer Radiation Dose Escalation Trial (n=870) | 60–80% | 1 / 2 (50.0%) | 0 / 2 (0.0%) |
|  | >80% | 1 / 1 (100.0%) | 0 / 1 (0.0%) |
| Prostate Cancer HRR Alteration Study (n=690) | 60–80% | 2 / 3 (66.7%) | 0 / 3 (0.0%) |
|  | >80% | 0 / 0 (—) | 0 / 0 (—) |
| Liver Cirrhosis & Hepatitis B Registry Study (n=667) | 60–80% | 9 / 19 (47.4%) | 0 / 19 (0.0%) |
|  | >80% | 2 / 7 (28.6%) | 0 / 7 (0.0%) |
| Recurrent Glioma Rhenium Nanoliposome Trial (n=608) | 60–80% | 3 / 5 (60.0%) | 0 / 5 (0.0%) |
|  | >80% | 0 / 0 (—) | 0 / 0 (—) |
| Chronic subdural hematoma embolization vs surgery study (n=381) | 60–80% | 0 / 0 (—) | 0 / 0 (—) |
|  | >80% | 0 / 3 (0.0%) | 0 / 3 (0.0%) |
| HER2-Positive Urothelial Cancer Immunotherapy Trial (n=381) | 60–80% | 0 / 2 (0.0%) | 0 / 2 (0.0%) |
|  | >80% | 0 / 0 (—) | 0 / 0 (—) |
| High-Risk Stage I NSCLC Surgery vs SABR Trial (n=257) | 60–80% | 0 / 1 (0.0%) | 0 / 1 (0.0%) |
|  | >80% | 0 / 0 (—) | 0 / 0 (—) |
| Leptomeningeal Metastases Prospective Study (n=157) | 60–80% | 0 / 0 (—) | 0 / 0 (—) |
|  | >80% | 0 / 0 (—) | 0 / 0 (—) |
| IDH1-Mutant Recurrent/Progressive Glioma Trial (n=154) | 60–80% | 0 / 0 (—) | 0 / 0 (—) |
|  | >80% | 0 / 0 (—) | 0 / 0 (—) |

**Table S5.** Representative per–criterion performance (selected). **Abbreviations:** Acc, accuracy; Sens, sensitivity; Spec, specificity; PPV, positive predictive value; NPV, negative predictive value; F1, F1-score; ER+, estrogen receptor–positive; SCC, squamous cell carcinoma; PET, positron emission tomography; PSMA-PET, prostate-specific membrane antigen PET; ADT, androgen deprivation therapy; BCR, biochemical recurrence; SSRI, selective serotonin reuptake inhibitor; SNRI, serotonin–norepinephrine reuptake inhibitor; UTI, urinary tract infection; *E. coli, Escherichia coli* ; Mets, metastases; N/A, not applicable (metric undefined due to zero denominator).

| Trial | Criterion | Acc | Sens | Spec | PPV | NPV | F1 |
| --- | --- | --- | --- | --- | --- | --- | --- |
| Calcium Phosphate Kidney Stone Study | STONE-FORMER | 0.991 | 1.000 | 0.500 | 0.991 | 1.000 | 0.996 |
| Calcium Phosphate Kidney Stone Study | NO-STRUVIITE | 1.000 | 1.000 | 1.000 | 1.000 | 1.000 | 1.000 |
| Early-Stage ER+ Breast Cancer Pre-Op Trial | TUMOR-DIMENSION | 1.000 | 1.000 | 1.000 | 1.000 | 1.000 | 1.000 |
| Early-Stage ER+ Breast Cancer Pre-Op Trial | NODE-NEGATIVE | 0.944 | 0.940 | 1.000 | 1.000 | 0.583 | 0.969 |
| Head & Neck SCC Radiation Trial | STAGING | 1.000 | 1.000 | 1.000 | 1.000 | 1.000 | 1.000 |
| High-Risk Prostate Cancer PET-Guided Radiotherapy Trial | PSMA-PET | 0.946 | 0.949 | 0.943 | 0.925 | 0.962 | 0.937 |
| Rectal Cancer Radiation Dose Escalation Trial | CIRCUMFERENCE | 1.000 | 1.000 | 1.000 | 1.000 | 1.000 | 1.000 |
| Rectal Cancer Radiation Dose Escalation Trial | SIZE | 1.000 | 1.000 | 1.000 | 1.000 | 1.000 | 1.000 |
| Liver Metastasis Ultra-High Dose Radiation Trial | LESION-SPECS | 0.766 | 0.727 | 0.806 | 0.800 | 0.735 | 0.762 |
| Liver Metastasis Ultra-High Dose Radiation Trial | LIVER-METS | 0.895 | 0.987 | 0.111 | 0.905 | 0.500 | 0.944 |
| Muscle-Invasive Urothelial Carcinoma Post-Surgery Trial | POST-SURGERY | 0.778 | 1.000 | 0.667 | 0.600 | 1.000 | 0.750 |
| Moderate-Severe Asthma Escitalopram Trial | EXACERBATIONS | 0.857 | 0.889 | 0.853 | 0.457 | 0.982 | 0.604 |
| Moderate-Severe Asthma Escitalopram Trial | NO-SSRI-SNRI | 0.949 | 1.000 | 0.385 | 0.947 | 1.000 | 0.973 |
| Prostate Cancer ADT Recurrence Study | TESTOSTERONE | 0.788 | 0.929 | 0.737 | 0.565 | 0.966 | 0.703 |
| Prostate Cancer ADT Recurrence Study | BCR | 0.857 | 0.923 | 0.706 | 0.878 | 0.800 | 0.900 |
| Prostate Cancer ADT Recurrence Study | PSMA-LESION | 1.000 | 1.000 | 1.000 | 1.000 | 1.000 | 1.000 |
| Drug-Resistant *E. coli* UTI Trial | ECOLI-RESIST | 0.826 | 1.000 | 0.000 | 0.826 | N/A | 0.905 |
| Drug-Resistant *E. coli* UTI Trial | NO-SULFA | 0.786 | 1.000 | 0.000 | 0.786 | N/A | 0.880 |

## Supplement B

### Standardized Setup Form

The AI team used an internal setup form to instantiate a trial configuration that parameterized the prescreening pipeline end-to-end. The form captured high-level variables only; no trial-specific logic was hard-coded beyond allowed filters or the criterion prompts. Provider rosters and criterion text were entered via the same interface but stored in separate tables.

### Parameter inventory (with examples)

- *Eligibility tiers and prioritization:* designation of the Tier 1 gate and a list of secondary criteria; optional priority for display triage (e.g., tumor size) without affecting logic.
- *Case-finding source and cadence:* suggested-provider lists or alternatives agreed with the study team (department panels, upcoming surgery schedules, modality-specific imaging orders); lookahead window and refresh cadence for prospective pulls.
- *Rescreening policy:* whether and when to rescreen previously reviewed patients; optional guidance to narrow subsequent retrievals.
- *Document scope and time windows:* trial-level look-back windows and inclusion filters for note types, laboratory tests, and imaging modalities; global exclusion of scanned/external PDFs by policy.
- *Retrieval and inference defaults:* embedding model, hierarchical parsing and node handling, target number of retrieved snippets (top-*K*), generator selection, compute routing, and run identifiers.
- *Database and security bindings:* read-only connections to institutional analytics warehouses; prospective pulls executed against analytics stores that typically lag the transactional source by approximately one day.
- *Paths to assets and outputs:* locations for prompt templates, criteria JSON, per-trial outputs, and update reports (environment-specific).
- *Notifications and contacts:* PI/research-staff contacts for status summaries and reminders; no PHI in notifications.

### Generating Eligibility Prompts

#### Prompt composition

Each criterion’s prompt comprised: (i) the criterion name; (ii) the verbatim protocol text; and (iii) a tips block that operationalized how retrieved evidence should be mapped to labels. Labels were: met, likely met, likely not met, not met, uncertain, no documents found. The tips block focused on clarity and reproducibility rather than exhaustively encoding protocol logic.

#### Decision rules for criterion labeling

The tips block used minimal, criterion-specific rules:

1. Assign *met* when explicit, in-window evidence satisfies all required conditions in the protocol text (e.g., disease identity/stage, laboratory thresholds, allowed imaging modality).
2. Assign *likely met* when evidence is strongly suggestive but missing a minor element (e.g., unit or date) or when multiple concordant notes imply the condition without a single definitive statement.
3. Assign *not met* when explicit, in-window evidence contradicts the criterion.
4. Assign *likely not met* when available evidence points against the criterion but remains incomplete or indirectly stated.
5. Assign *uncertain* when evidence is internally conflicting, temporally ambiguous, or insufficient to support a directional judgment.
6. Assign *no documents found* when retrieval returns no relevant evidence after filtering and similarity search; absence of documentation alone is not used to infer failure.

#### Literature context from PubMed

A companion agent transformed the free-text criterion into a structured PubMed query and retrieved recent review literature (titles/abstracts; open-access full text when available). The agent summarized definitional boundaries, common synonyms/abbreviations, and typical temporal qualifiers into a brief context paragraph that was appended to the tips block prior to inference. Only the criterion text was sent to PubMed; no patient data were transmitted. Pseudocode for prompt generation can be found in Supplement C: Initial literature-assisted tip generation.

## Supplement C

### Tiered prescreen processing

At each run, the system loads past results, gathers the upcoming patient list plus any cases scheduled for recheck, and reads the trial rules grouped as Tier 1 (must-have), Tier 2, and optional Tier 3. It reviews patients one by one, clearing only the model’s prior outputs for rechecks while preserving human notes. Tier 1 is evaluated first and acts as a gate: if labeled *not met, likely not met, uncertain*, or *no documents found*, lower tiers are skipped. When Tier 1 is *met* or *likely met*, the system proceeds to Tier 2 and, if set, Tier 3. For each rule, it records a structured answer (status, confidence, brief explanation, tier) and saves the supporting text with document dates for audit. After each patient, results are saved, the review queue is reprioritized, and the recheck list is updated. **Disclaimer: pseudocode was AI-generated using source code**.

### Pseudocode

~~~
FUNCTION OrchestrateTieredPrescreen(cfg, criteria_json, providers_json,
                                     input_dir, prompt_template, output_csv,
                                     overwrite = FALSE, evaluation_mode = “cot”):
       # 0) Initialize and validate required parameters
       REQUIRE cfg has {tier1, tier2, process_tier3, priority_column,
                     database, server, port, table_name, index_column, driver}
       start_time <- NOW()
       # 1) Load or initialize outcomes table (CSV)
       outcomes <- LoadCSVOrInit(output_csv, index_col = cfg.index_column)
       # 2) Fetch prospective roster and rescreen ledger from SQL
       roster_ids <- SQL_GetRosterIDs(cfg.database, cfg.server, cfg.port,
                                            cfg.table_name, cfg.index_column, cfg.driver) rescreen_ids <- SQL_GetRescreenList(cfg)
       # 3) Load and organize criteria
       C <- LoadCriteria(criteria_json)          # ordered dict
       tier1 <- cfg.tier1                        # list
       tier2 <- cfg.tier2                        # list
       tier3 <- (AllKeys(C) \ (tier1 tier2)) if cfg.process_tier3 else []
       EnsureOutcomeColumns(outcomes, C, cfg.index_column)
       # 4) Iterate through patient staging folders (one folder per patient)
       FOR each folder_name IN ListDirs(input_dir):
           patient_id <- ParseIntOrSkip(folder_name)
           IF NOT ShouldProcess(patient_id, roster_ids, outcomes,
                                priority = cfg.priority_column,
                                overwrite = overwrite, cfg = cfg):
                 CONTINUE
           # 4a) Rescreen handling: remove prior AI artifacts, preserve human review
           IF patient_id IN rescreen_ids:
                 SQL_RemoveAIArtifacts(patient_id, cfg) # e.g., “-TS”, “-source” tables
           # 4b) Ensure patient row exists
           outcomes <- EnsureRow(outcomes, cfg.index_column, patient_id)
           # 5) Evaluate against tiers
           outcomes <- EvaluatePatientAgainstTiers(patient_id, folder_name,
                                                         C, tier1, tier2, tier3,
                                                         prompt_template, evaluation_mode,
                                                         outcomes, cfg)
           # 6) Persist after each patient
           SaveCSV(outcomes, output_csv)
           ReorderForReview(output_csv, priority_col = cfg.priority_column)
           SQL_WriteDenormalized(output_csv_reordered, cfg)
           SQL_UpdateRescreenLedger(outcomes, cfg)
       LogRunSummary(start_time, output_csv)
       RETURN outcomes
FUNCTION EvaluatePatientAgainstTiers(patient_id, patient_dir,
                                     C, tier1, tier2, tier3,
                                     prompt_template, evaluation_mode,
                                     outcomes, cfg):
       # Context manager preps patient context, retrieval, and prompting
       WITH BuildTierContext(patient_id, patient_dir, C, prompt_template,
                             evaluation_mode, cfg) AS state:
          # Tier 1 gate
          FOR crit IN tier1:
              outcomes, failed_name, tier1_failed, model_error <-
                   EvaluateSingleCriterion(patient_id, crit, state, outcomes, cfg)
              IF model_error:
                   HandleModelFailure(state, patient_id, cfg)
                   BREAK
              IF tier1_failed:
                   Log(“Tier1 failed on” + failed_name)
                   BREAK
          # If Tier 1 failed, mark remaining tiers as not evaluated with reason
          IF tier1_failed:
              outcomes <- MarkNotEvaluated(patient_id, tier2 tier3,
                                                 reason = “Tier1 gate not satisfied”,
                                                 state, outcomes, cfg)
              RETURN outcomes
          # Tier 2 block
          FOR crit IN tier2:
              TRY:
                   outcomes, _, _, model_error <-
                        EvaluateSingleCriterion(patient_id, crit, state, outcomes, cfg)
              CATCH e:
                   outcomes <- MarkNotEvaluated(patient_id, [crit],
                               reason = “Internal AI error at tier 2”, state, outcomes, cfg)
          # Tier 3 block (optional)
          IF cfg.process_tier3:
               FOR crit IN tier3:
                  TRY:
                      outcomes, _, _, model_error <-
                            EvaluateSingleCriterion(patient_id, crit, state, outcomes, cfg)
                  CATCH e:
                      outcomes <- MarkNotEvaluated(patient_id, [crit],
                            reason = “Internal AI error at tier 3”, state, outcomes, cfg)
          ELSE:
               outcomes <- MarkNotEvaluated(patient_id, tier3,
                                                  reason = “process_tier3 is False”,
                                                  state, outcomes, cfg)
          RETURN outcomes
FUNCTION EvaluateSingleCriterion(patient_id, criterion_name, state, outcomes, cfg):
    # Fail statuses gate later tiers; matches paper definitions
    fail_statuses <- {“not met”, “likely not met”, “uncertain”, “no documents found”}
    WITH BuildCriterionContext(patient_id, criterion_name, state,
                               evaluation_mode = state.evaluation_mode,
                               fail_statuses = fail_statuses, cfg = cfg) AS ctx:
        # Run retrieval + prompting and parse structured JSON
        model_payload <- ctx.RunModel()
        resp <- ParseJSONWithMarkdownFallback(model_payload[criterion_name].response)
        # Assign tier for provenance if missing
        IF NOT HasKey(resp, “tier”):
            resp[“tier”] <- InferTier(criterion_name, state.tier_map)
        # Update outcomes table with status, certainty, support, free text, tier
        outcomes <- UpdateOutcomes(outcomes, patient_id, criterion_name, resp, state)
        # Persist sources: normalized and raw
        src_text <- model_payload[criterion_name].source_text
        IF src_text IS NOT EMPTY:
              SaveRawSource(patient_id, criterion_name, src_text, cfg)
              norm_text, doc_date <- NormalizeSource(src_text, index_dir = state.index_dir)
              SQL_WriteSource(patient_id, criterion_name, doc_date, norm_text, cfg, table_suffix=“-
              SQL_WriteSource(patient_id, criterion_name, doc_date, src_text, cfg, table_suffix=“-
        # Determine gating outcome
        IF resp.status IN fail_statuses:
           RETURN outcomes, criterion_name, TRUE, FALSE # failed Tier gate
        ELSE:
           RETURN outcomes, NULL, FALSE, FALSE          # passed
~~~

### Pseudocode

~~~
FUNCTION GenerateInitialTipsFromLiterature(criterion_text,
                                           seed_tips = ““,
                                           retmax = 10,
                                           article_type = “[Review]”,
                                           language = “English”,
                                           model_for_query = “gpt-4.1”,
                                           model_for_tips = “gpt-4.1”):
         # 1) Build a structured PubMed query from the criterion text
         query_prompt <- BuildPubMedQueryPrompt(criterion_text)
         SetModel(model_for_query)
         query_obj <- LLM_StructuredOutput(prompt = query_prompt,
                                                 schema = PubMedQueryModel)
         refined_query <- Trim(query_obj.refined_query)
         # 2) Retrieve recent review literature and construct a small index
         pm_records <- PubMedFetch(query = refined_query,
                                         retmax = retmax,
                                         restrict_to_reviews = TRUE,
                                         free_full_text_preferred = TRUE,
                                         language = language)
         docs <- LoadPubMedRecords(pm_records)         # titles/abstracts; OA full text if available
         lit_index <- BuildVectorIndex(docs, embedding_model = “text-embedding-3-large”)
         # 3) Extract clarifying context from the literature index
         context_prompt <- BuildContextExtractionPrompt(criterion_text, seed_tips)
         context_result <- lit_index.Query(context_prompt)
         context_info <- String(context_result.response)
         source_citations <- ConcatenateSourceDetails(context_result.source_nodes) # for logging
         # 4) Synthesize a COMPLETE decision-tree “tips” block
         tips_prompt <- BuildCriterionTipsPrompt(criterion = criterion_text,
                                                       current_tips = seed_tips,
                                                       context_info = context_info)
         SetModel(model_for_tips)
         tips_candidate <- LLM_Chat(tips_prompt)
         # 5) Return proposed tips and log provenance (no patient data shared)
         AuditLog.Write({
            “query”: refined_query,
            “pmids”: pm_records.PMIDs(),
            “article_type”: article_type,
            “language”: language,
            “model_for_query”: model_for_query,
            “model_for_tips”: model_for_tips,
            “timestamp”: NOW()
         })
         RETURN tips_candidate
~~~

### Human-in-the-loop, automated prompt refinement

We combined routine feedback with a light-touch automation step to keep *tips* accurate over time. During the first month of a trial, we met every two weeks, then monthly, to review criterion-level comments from study staff. Each comment was labeled by outcome (false/true positive or negative) and mapped to a simple error taxonomy (boundary/threshold, terminology/synonym, temporal qualifier, document provenance). When the same issue recurred (e.g., two or more times) or a reviewer explicitly requested a change, a refinement agent produced a full replacement tips block that preserved correct rules and edited only what was needed. When enabled, the agent could also consult the literature by turning the criterion into a structured PubMed query that favored recent reviews, indexing titles/abstracts (and open-access full text when available), and extracting clarifying context. Only the criterion text and de-identified reviewer summaries were used for this step; no patient data were shared. Candidates were presented in the internal UI for human approval. Approved changes were versioned and logged, and became active at the next run; historical AI outputs and human labels were not altered.**Disclaimer: pseudocode was AI-generated using source code**.

### Pseudocode

~~~
FUNCTION ProposeTipRefinement(criterion_name,
                              criterion_text,
                              current_tips,
                              reviewer_comments_dict,
                              use_literature = TRUE,
                              k_reviews = 5):
         # 1) Validate inputs
         REQUIRE criterion_text, current_tips, reviewer_comments_dict
         # 2) Summarize reviewer feedback by error class
            # reviewer_comments_dict keys: {FP, FN, TP, TN}
            feedback_digest <- JoinNonEmpty(reviewer_comments_dict, sep=“\n”)
            literature_context <- ““
            IF use_literature == TRUE:
                # 3) Build a structured PubMed query from the criterion text
                pm_query_object <- LLM_StructuredQuery(
                     prompt = BuildPubMedQueryPrompt(criterion_text, feedback_digest
                )
                # 4) Retrieve recent reviews and build a small literature index
                pm_records <- PubMedFetch(query = pm_query_object.query,
                                                retmax = k_reviews,
                                                restrict_to_reviews = TRUE,
                                                language = “English”,
                                                free_full_text_preferred = TRUE)
                lit_index <- BuildVectorIndex(pm_records) # titles/abstracts, open access full text
                # 5) Extract clarifying context for terms, thresholds, time windows
                literature_context <- lit_index.Query(
                    BuildContextExtractionPrompt(criterion_name, current_tips)
                )
            # 6) Ask an LLM to synthesize a COMPLETE, revised tips block
            #   preserving unchanged rules and editing only what feedback supports
            refined_tips <- LLM_Generate(
               prompt = BuildTipRefinementPrompt(
                           criterion_text = criterion_text,
                           current_tips = current_tips,
                           feedback_digest = feedback_digest,
                           literature_context = literature_context,
                           guardrails = “Edit only if FP/FN show a systematic issue;
                                        otherwise return current_tips verbatim.”)
            )
            RETURN refined_tips
FUNCTION SubmitForApprovalAndVersioning(criterion_id, refined_tips, active_flag):
            # 7) Present candidate tips in the internal UI for human review
            approved <- HumanApproverConfirms(criterion_id, refined_tips)
            IF approved:
                # 8) Archive previous tips, activate new tips prospectively
                prior_tips <- DB.Read(StudyCriteria[criterion_id].tips)
                DB.Update(StudyCriteria[criterion_id],
                          tips = refined_tips,
                          change_from_prior = prior_tips,
                          active = active_flag,
                          updated_at = NOW())
                # 9) Log provenance (cfg hash, model/embedding versions, PMIDs, timestamps)
                AuditLog.Write({
                     “criterion_id”: criterion_id,
                     “action”: “tips_update”,
                     “pmids”: pm_records.PMIDs if use_literature else [],
                     “model_version”: CurrentModel(),
                     “embedding_version”: CurrentEmbeddingModel(),
                     “cfg_hash”: CurrentCfgHash(),
                     “timestamp”: NOW()
                })
                RETURN “updated”
            ELSE:
                RETURN “rejected”
         # Trigger conditions (examples)
         IF RecurrentErrorCount(criterion_id, class=“FP” or “FN”) >= 2             OR ReviewerRequestsChange(criterion_id):
              candidate <- ProposeTipRefinement(…)
              SubmitForApprovalAndVersioning(criterion_id, candidate, active_flag=TRUE)
         # Note: No retrospective re-scoring is performed.
~~~

